# SARS-CoV-2 antibody landscape following multiple vaccinations and infections during Omicron period

**DOI:** 10.64898/2026.11.16.26368199

**Authors:** Yangyupei Yang, Casey Juntila-Raymond, Amy Callear, Matthew Smith, Elie-Tino Godonou, Arnold S. Monto, Emily T. Martin

**Affiliations:** Department of Epidemiology, University of Michigan School of Public Health, Ann Arbor, MI, USA

**Author notes:** **Corresponding author:** Yangyupei Yang, Emily T. Martin.

## Abstract

**Background:** Repeated COVID-19 vaccination effects on the immunogenicity, breadth, and durability of antibody responses across antigenically distinct SARS-CoV-2 variants remain incompletely understood.

**Method:** Participants receiving ≥five COVID-19 vaccine doses 2020-2025 were selected from a prospective cohort in Michigan. Serum IgG, IgM, and ACE2 percent inhibition were measured using multiplex assays. Baseline-adjusted immunogenicity, cross-reactivity, and formulation-specific antibody durability were evaluated using regression models.

**Result:** Twenty-one participants were included (median: seven vaccine doses). For every two-fold increase in pre-vaccination IgG concentration, post-vaccination fold-rise was 34% (95% CI: 16%–48%), 43% (95% CI: 15%–62%), and 55% (95% CI: 21%–75%) lower following monovalent, bivalent, and 2023– 24 vaccination, while baseline-adjusted immunogenicity showed no significant trends across successive vaccines. Antibody landscapes demonstrated broad cross-reactivity against ancestral and Omicron variants, with similar strain-specific IgG changes across ancestral, vaccine-matched, and antigenically distant strains, but greater heterogeneity in ACE2 inhibition. Formulation-specific waning models showed significant IgG declines following monovalent and bivalent vaccination (β = −1.03, 95% CI: −1.51 to −0.55; P<0.001 and −1.00, 95% CI: −1.71 to −0.30; P=0.007, respectively) and ACE2 inhibition declines following monovalent vaccination, while other formulations showed less consistent changes. Despite waning, antibody levels generally remained relatively high across variants up to 300 days post-vaccination, with formulation-specific differences in durability.

**Conclusion:** Repeated COVID-19 vaccination maintained baseline-adjusted immunogenicity and broad cross-reactive antibody responses, with formulation-specific differences in durability. Higher baseline immunity was associated with smaller post-vaccination increases, suggesting repeated boosting continues to reinforce humoral immunity despite increasingly complex immune histories.

## Introduction

The Continued evolution of SARS-CoV-2 and the emergence of antigenically distinct Omicron subvariants have resulted in recommendations for revaccination with updated COVID-19 vaccine formulations in order to maintain vaccine-induced immunity.^1,2,3(pp2024-2025),4^. Many individuals have now received multiple sequential COVID-19 booster doses, creating increasingly complex immune histories shaped by repeated vaccination, prior infection, and exposure to antigenically distinct viral variants^5–8^. Understanding how repeated vaccination influences the magnitude, breadth, and durability of antibody responses to subsequent doses has become an important consideration for future vaccine strain selection and vaccination strategies^9^.

Repeated exposure to antigenically distinct vaccine antigens may not only enhance responses against the vaccine strain but also recall immunity generated by previous infections or vaccinations^10–12^. Prior studies have shown that vaccination or infection can induce antibody responses not only against contemporary strains but also against previously encountered antigenically related variants, commonly referred to as “backboosting”^10,13^. The role of backboosting following repeated COVID-19 vaccination remains incompletely understood. Determining whether repeated booster vaccination broadens cross-reactive immunity across historical and newly emerging SARS-CoV-2 variants may provide insight into how prior immune history shapes responses to updated vaccines. Higher pre-vaccination antibody levels have been associated with smaller post-vaccination antibody increases, but it remains unclear whether these attenuated responses reflect antibody ceiling effects or reduced immune responsiveness following repeated vaccination^6^. Furthermore, whether repeated vaccination maintains functional antibody activity and durable immunity despite progressively higher baseline antibody levels has not been well characterized^14–17^.

Using longitudinal serological data from highly vaccinated participants in the Household Influenza Vaccine Evaluation (HIVE) cohort, we evaluated binding and functional antibody responses following up to seven COVID-19 vaccine doses administered between 2020 and 2025. Specifically, we evaluated booster immunogenicity after accounting for baseline antibody levels, characterized cross-reactive antibody responses across antigenically distinct SARS-CoV-2 variants to assess evidence of backboosting, and assessed the durability of antibody responses following repeated booster vaccination.

## Methods

### 2.1 Data source

We used data from the Household Influenza Vaccine Evaluation (HIVE), a prospective cohort study that conducted active acute respiratory illness (ARI) surveillance on 300-400 households annually since 2011^18^. Participant data included demographic and household characteristics, symptomatic respiratory infections including SARS-CoV-2 infection, and biannual as well as pre- and post-vaccination serological samples. COVID-19 vaccination details of study participants were obtained from the Michigan state immunization information system, electronic medical records, and self-reports through surveys^18^.

The analysis included participants enrolled in the HIVE cohort between August 2020 and August 2025, who had received at least five doses of mRNA COVID-19 vaccine, and had at least one post-vaccination serological sample collected following a COVID-19 booster dose during the study period. Vaccinations were classified into four formulation groups: original monovalent, bivalent, 2023–24 updated, and 2024– 25 updated formulations (Table S1).

### 2.2 Lab methods

Serological samples were tested for IgG and IgM antibody concentrations and ACE2 percentage inhibition using V-PLEX SARS-CoV-2 panels 32 for IgG, IgM, and panel 33 and 38 for ACE2 ^19,20(p32),21(p33)^ (Meso Scale Discovery; Table S2) following the manufacturer’s protocol^22^ and previously described elsewhere^6,7^. To detect SARS-CoV-2 infections, nasal swabs collected from participants who reported two or more ARI symptoms were tested for SARS-CoV-2 virus.

### 2.3 Statistical Analysis

Analyses were conducted according to COVID-19 booster vaccine formulation. For each vaccine formulation, we selected the corresponding vaccine antigens to evaluate antibody responses and durability following vaccination. Participant demographic characteristics, household characteristics, vaccination history, and SARS-CoV-2 infection history were summarized descriptively across the study population. Additional details on vaccine formulation classification, formulation-specific antigen selection, antibody measures, serum selection, and statistical models are provided in the Supplementary Methods.

#### 2.3.1 Repeated booster vaccine immunogenicity

We evaluated IgG and IgM geometric mean concentrations (GMCs), and mean ACE2 percent inhibition before and after vaccination across four booster vaccine formulation groups. Peak post-vaccination responses were assessed using samples collected 10-45 days after vaccination (closest to day 15 if multiple post vaccination samples), with the peak post-vaccination response defined as the sample collected closest to day 15 after vaccination. For participants with paired pre- and post-vaccination samples for each booster dose, we calculated IgG and IgM geometric mean fold-rises (GMFRs), and mean changes in ACE2 percent inhibition.

To evaluate the relationship between baseline antibody levels and post-vaccination boosting, we used linear regression models with pre-vaccination antibody measurements as predictors of subsequent antibody responses. We further evaluated baseline-adjusted immunogenicity using residuals from these models. Vaccine formulation was coded as an ordinal variable order by year of formulation licensure, and linear regression was used to assess trends in immunogenicity scores across successive formulations. Detailed model specifications are provided in the Supplementary Methods.

#### 2.3.2 Antibody Landscapes

We evaluated the breadth and magnitude of antibody responses across SARS-CoV-2 variants using two complementary approaches. First, descriptive antibody landscapes were constructed to visualize IgG and IgM GMCs and mean ACE2 percent inhibition before and after vaccination with each vaccine formulation, with SARS-CoV-2 antigens arranged chronologically according to their emergence. Additional landscapes compared peak post-vaccination responses with later follow-up responses to characterize antibody durability across variants.

In the second approach, we constructed antibody landscapes based on antigenic cartography. Published neutralization data reported by Rössler et al^11^ were used to reconstruct a two-dimensional antigenic map (Figure S1). IgG antibody concentrations were projected onto this map to visualize the magnitude and breadth of antibody responses across antigenic space following different vaccine formulations. Details of antigenic distance estimation and landscape construction are provided in the Supplementary Methods.

#### 2.3.3 Antibody Durability

Antibody durability was evaluated using samples collected 10-300 days after vaccination, grouped by vaccine formulation. The pre-next-vaccination response was defined as the latest available sample collected before the next vaccination date and at least 60 days after the peak sample. Descriptive analyses compared peak post-vaccination responses with later measurements collected before subsequent vaccination, including IgG and IgM GMCs, GMFCs, and ACE2 mean percent inhibition. Power-law models were fitted for each vaccine formulation and antibody target to characterize changes in antibody levels with time since vaccination. Model-predicted antibody levels were estimated at 15, 180, and 300 days post-vaccination. Model specifications, sampling restrictions, and sensitivity analyses are described in the Supplementary Methods.

Sensitivity analyses included leave-one-out cross-validation (LOOCV) at the participant level. All analyses were conducted using R (version 4.5.0).

## Results

Overall, 21 HIVE participants were included for this analysis, with a mean age of 51 years (IQR: 50, 56), and median of 7 (range: 5–9) COVID-19 vaccine doses received during the study period (Figure S2, Table S3). Booster vaccinations were categorized into four formulation groups: original monovalent, bivalent, 2023–24, and 2024–25. Across booster doses, the proportion of participants with prior SARS-CoV-2 infection before vaccination was 15.8%, 42.9%, 23.1%, and 66.7%, respectively (Figure 1).

**Figure 1.**
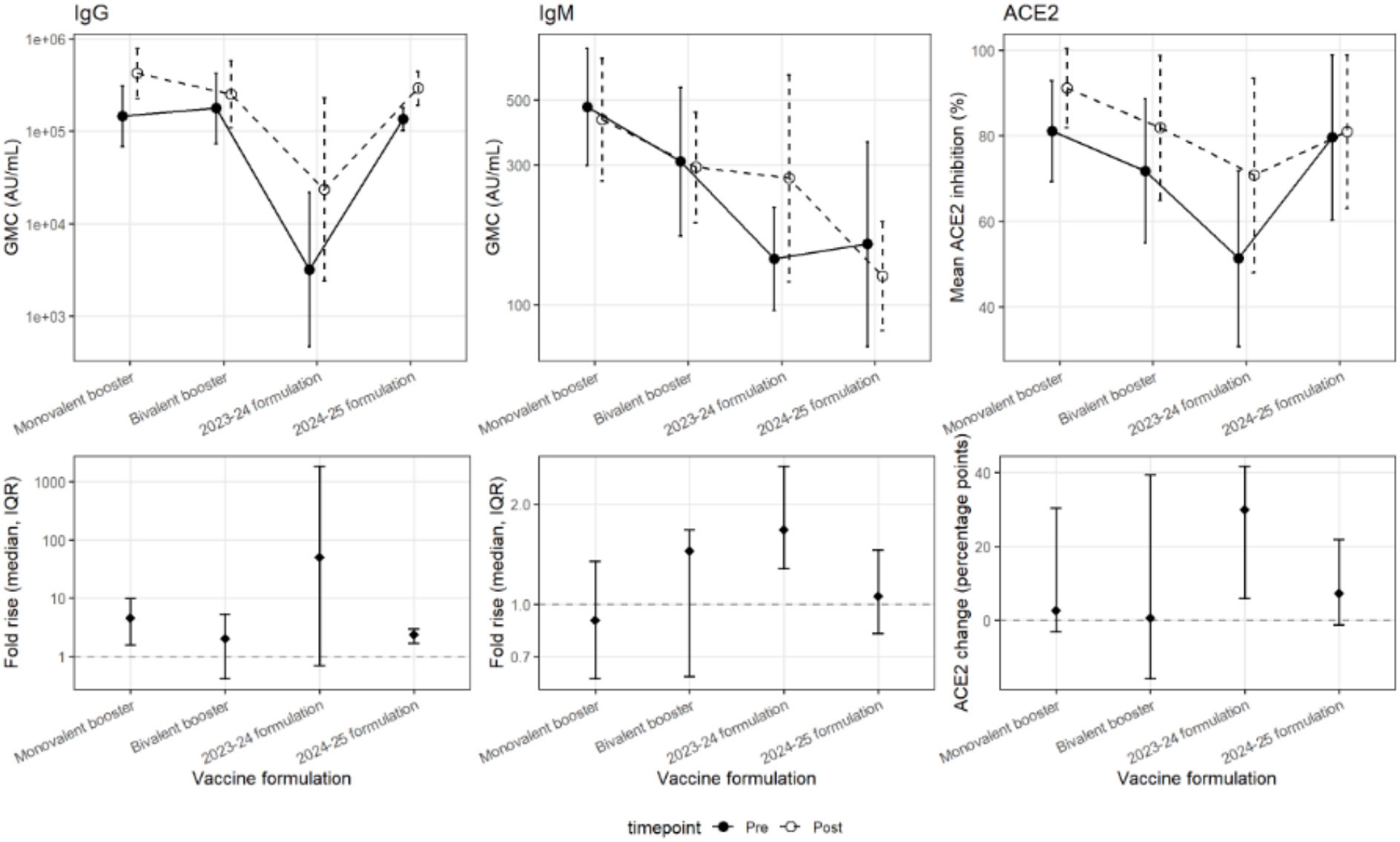
Antibody responses before and after COVID-19 booster vaccination by vaccine formulation. IgG, IgM, and ACE2 inhibition responses are shown across four vaccine formulations: original monovalent, bivalent, 2023–2024, and 2024–2025. For IgG and IgM, points and lines represent geometric mean concentrations with 95% confidence intervals. For ACE2, points and lines represent mean percent inhibition with 95% confidence intervals. Pre-vaccination responses are shown with solid lines and filled circles, and post-vaccination responses with dashed lines and open circles. Lower panels summarize the median post-vaccination fold-rise with interquartile range for IgG and IgM and the median post-minus pre-vaccination change in ACE2 percent inhibition with interquartile range. Each eligible vaccination event was analyzed separately, including multiple vaccination events from the same participant.

### 3.1 Antibody response by vaccine booster dose

IgG concentrations increased following vaccination across all four formulations, although the magnitude of boosting varied (Figure 1, Table S4). Pre- and post-vaccination IgG geometric mean concentrations (GMCs) were 146,010 and 423,570 AU/mL following monovalent vaccination, compared with 177,160 and 252,232 AU/mL following bivalent vaccination. The corresponding GMCs were 3,197 and 23,459 AU/mL for the 2023–24 formulation and 136,036 and 291,974 AU/mL for the 2024–25 formulation. Median IgG fold-rises were 4.5, 2.0, 50.6, and 2.4, respectively, with substantial variability in the 2023– 24 group. ACE2 inhibition generally increased following vaccination, whereas IgM responses were smaller and less consistent across formulations.

### 3.2 Repeated vaccination immunogenicity

Higher pre-vaccination antibody levels were generally associated with smaller post-vaccination responses across booster doses compared to individuals with lower pre-vaccination levels(Figure 2, Table S5). For every two-fold increase in baseline IgG concentration, post-vaccination fold-rise was approximately 34%, 43%, and 55% lower following monovalent (β = −0.59, 95% CI: −0.94 to −0.25; P = 0.003), bivalent (β = −0.81, 95% CI: −1.40 to −0.23; P = 0.01), and 2023–24 vaccination (β = −1.16, 95% CI: −1.98 to −0.34; P = 0.01), respectively. No statistically significant association was observed following the 2024–25 formulation. Similar inverse associations were observed for ACE2 inhibition following monovalent and bivalent vaccination, whereas associations following the 2023–24 and 2024–25 formulations were not statistically significant. For IgM, significant inverse associations were observed following bivalent and 2024–25 vaccination, with less consistent associations for other formulations.

**Figure 2.**
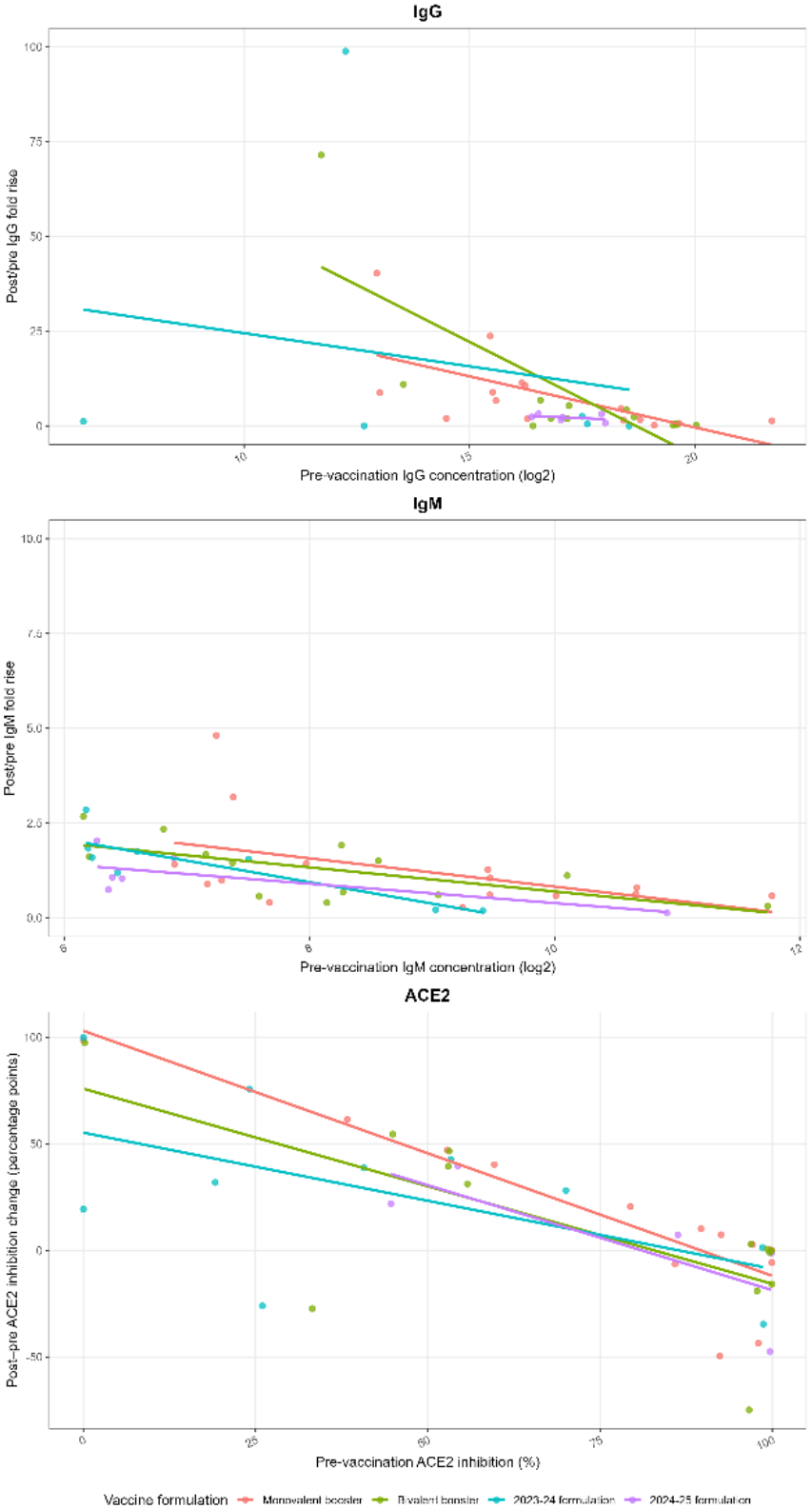
Association between pre-vaccination antibody levels and post-vaccination antibody responses by COVID-19 booster vaccination formulation. Relationships between pre-vaccination antibody levels and post-vaccination responses are shown by vaccine formulation for (A) IgG, (B)IgM, (C) ACE2 percent inhibition. For IgG and IgM, the y-axis represents the fold-rise between pre- and post-vaccination samples. For ACE2, the y-axis represents the absolute change in percent inhibition following vaccination. Colored points represent individual vaccination events, and separate formulation-specific linear regression lines were fitted for the original monovalent, bivalent, 2023–2024, and 2024–2025 formulations. Regression lines are displayed only across the observed range of pre-vaccination antibody levels for each formulation.

After accounting for pre-vaccination antibody levels, we found no statistically significant changes in immunogenicity across four chronologically ordered vaccine formulations (Table S6, Figure S3). After adjusting for pre-vaccination antibody levels, no significant changes in immunogenicity were observed with each additional vaccine dose for IgG (β = 1.22, 95% CI: −3.27 to 5.71; P = 0.57), IgM (β = −0.00, 95% CI: −0.27 to 0.27; P = 0.98), or ACE2 inhibition (β = −0.25, 95% CI: −11.28 to 10.79; P = 0.96). These findings suggest no clear evidence of progressively reduced immunogenicity following repeated COVID-19 vaccination after accounting for baseline antibody levels.

Leave-one-participant-out cross-validation sensitivity analyses showed generally consistent patterns and directions of association compared with the primary analyses (Table SX).

### 3.3 Antibody Landscapes

Antigen-specific analyses demonstrated broad IgG cross-reactivity against ancestral and Omicron SARS-CoV-2 variants following vaccination with different formulations (Figure 3, Table S7). Antibody concentrations varied across antigens and formulations, with generally higher IgG responses against the ancestral strain and BA.4/5-related variants than against antigenically distant XBB-related variants. Strong boosting against vaccine-matched antigens were observed following each formulation, including ancestral strain responses after monovalent vaccination, ancestral and BA.4/5 responses after bivalent vaccination, XBB.1.5-related responses after 2023–24 vaccination, and JN.1-related responses after 2024–25 vaccination. These responses were accompanied by varying degrees of cross-reactivity against antigens not included in the corresponding formulations, and backboosting of antibody responses against strains targeted by previous vaccine formulations. IgM responses were smaller and more heterogeneous, while ACE2 inhibition generally increased after vaccination, with variation across antigen targets.

**Figure 3.**
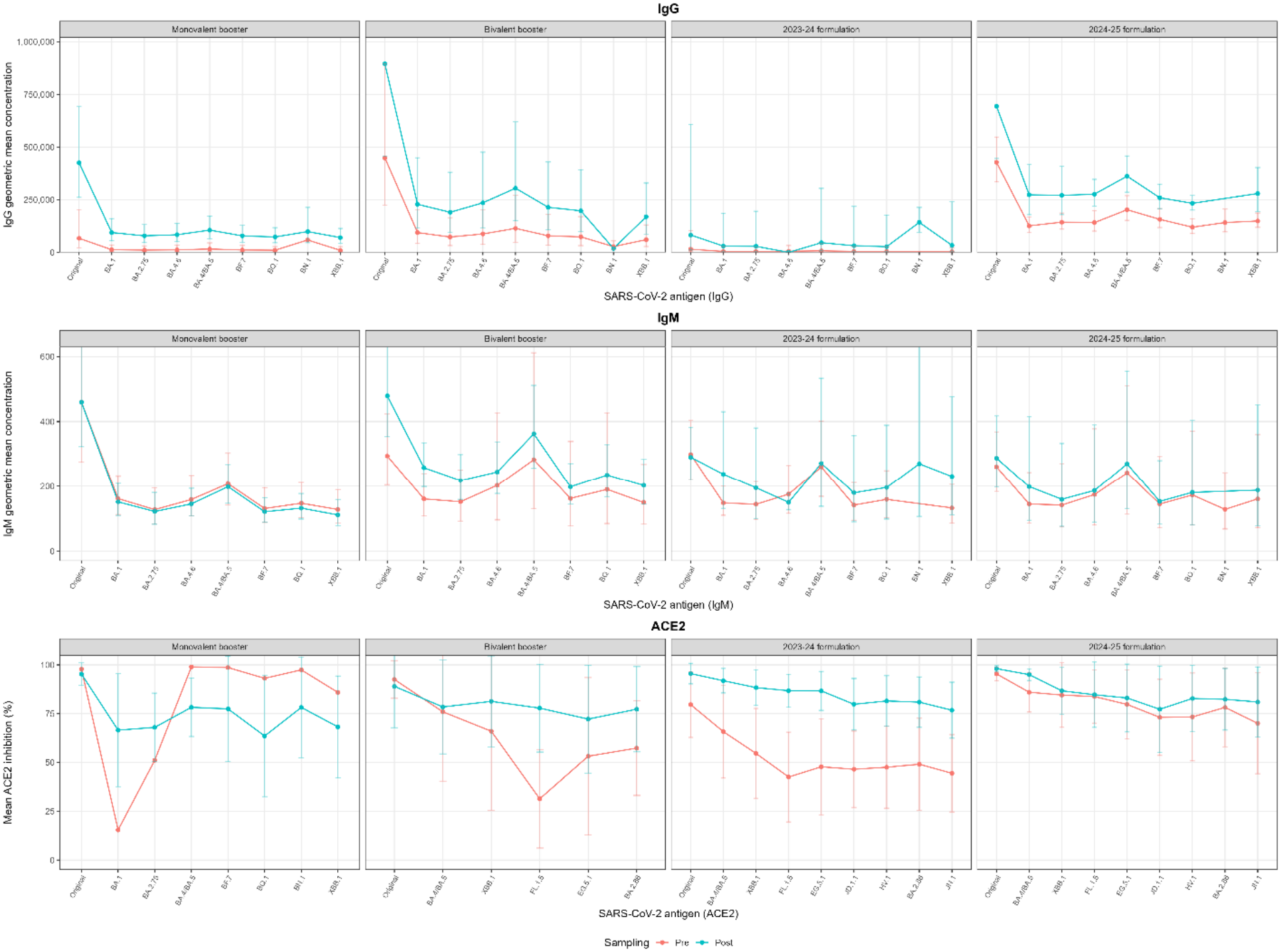
Antibody responses before and after COVID-19 booster vaccination across SARS-CoV-2 variants by vaccine formulation. Antibody responses before (Pre) and after (Post) COVID-19 booster vaccination are shown for (A) IgG, (B) IgM, and (C) ACE2 percent inhibition across SARS-CoV-2 variants. Panels are presented separately for the original monovalent, bivalent, 2023–2024, and 2024–2025 vaccine formulations. Pre-vaccination samples were selected as the closest available serum samples collected before the current vaccination and at least 90 days after the preceding vaccination, when applicable. Lines represent geometric mean concentrations for IgG and IgM or mean percent inhibition for ACE2.

IgG antibody landscapes constructed using a previously published antigenic map showed board reactivity across antigenic space following repeated vaccination ^12^(Figure S1, Figure 4). Antibody responses against Omicron variants, including BA.5, BQ.1.1, and XBB.1, also increased following repeated vaccination, with broader and higher post-vaccination reactivity observed after the multiple booster dose compared with the first monovalent booster dose.

**Figure 4.**
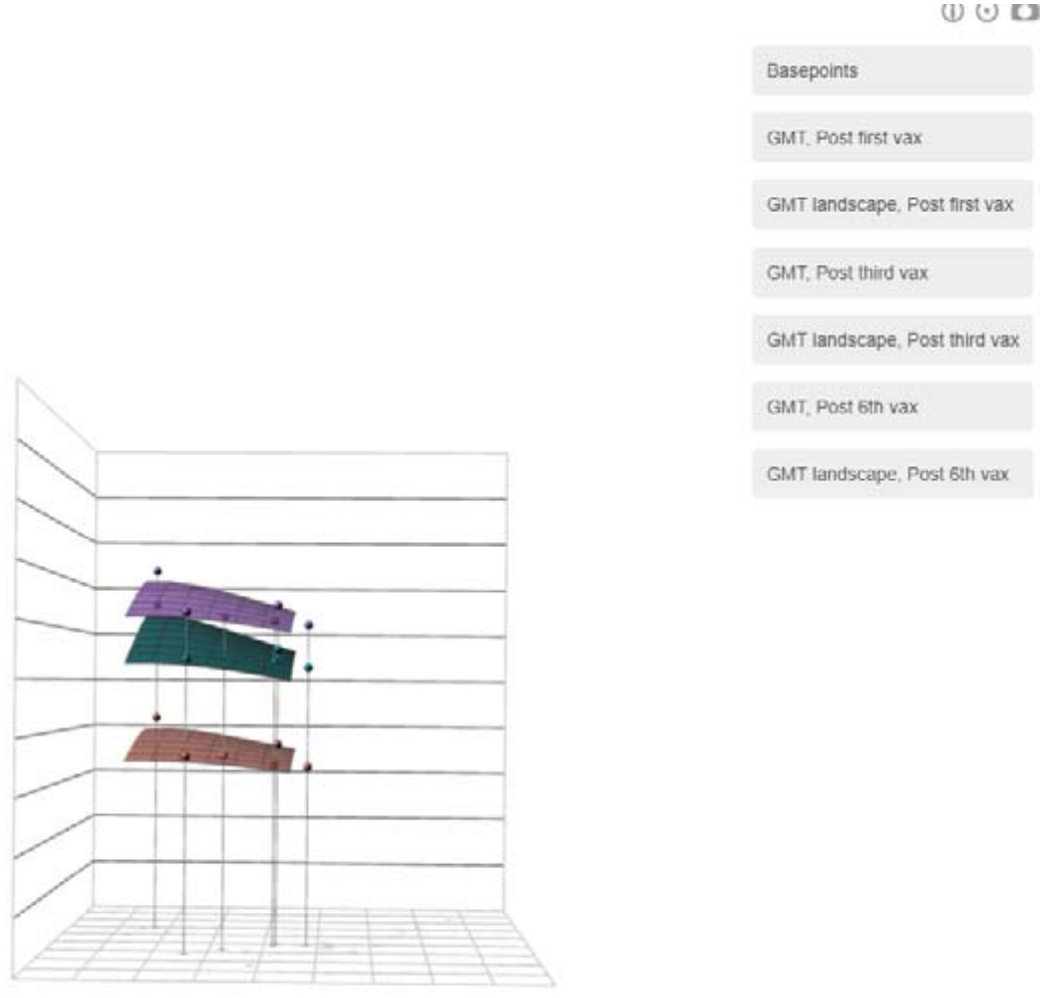
Antibody landscapes following COVID-19 vaccination.Antibody landscapes showing IgG responses following vaccination with primary dose, monovalent booster, and 2024–2025 COVID-19 vaccine formulations, projected onto a published SARS-CoV-2 antigenic map (ref). The x- and y-axes represent antigenic space derived from published antigenic cartography, while the z-axis represents log2-transformed IgG concentrations. The landscapes illustrate differences in the magnitude and breadth of antibody responses across vaccine formulations. The underlying SARS-CoV-2 antigenic map used to construct the antibody landscapes is shown in Supplementary Figure S1.

### 3.4 Antibody Durability

Antibody responses generally declined following vaccination, although the magnitude and statistical evidence for waning differed across vaccine formulations and antibody measurements (Figures 5–6, Tables S8–S10).

**Figure 5.**
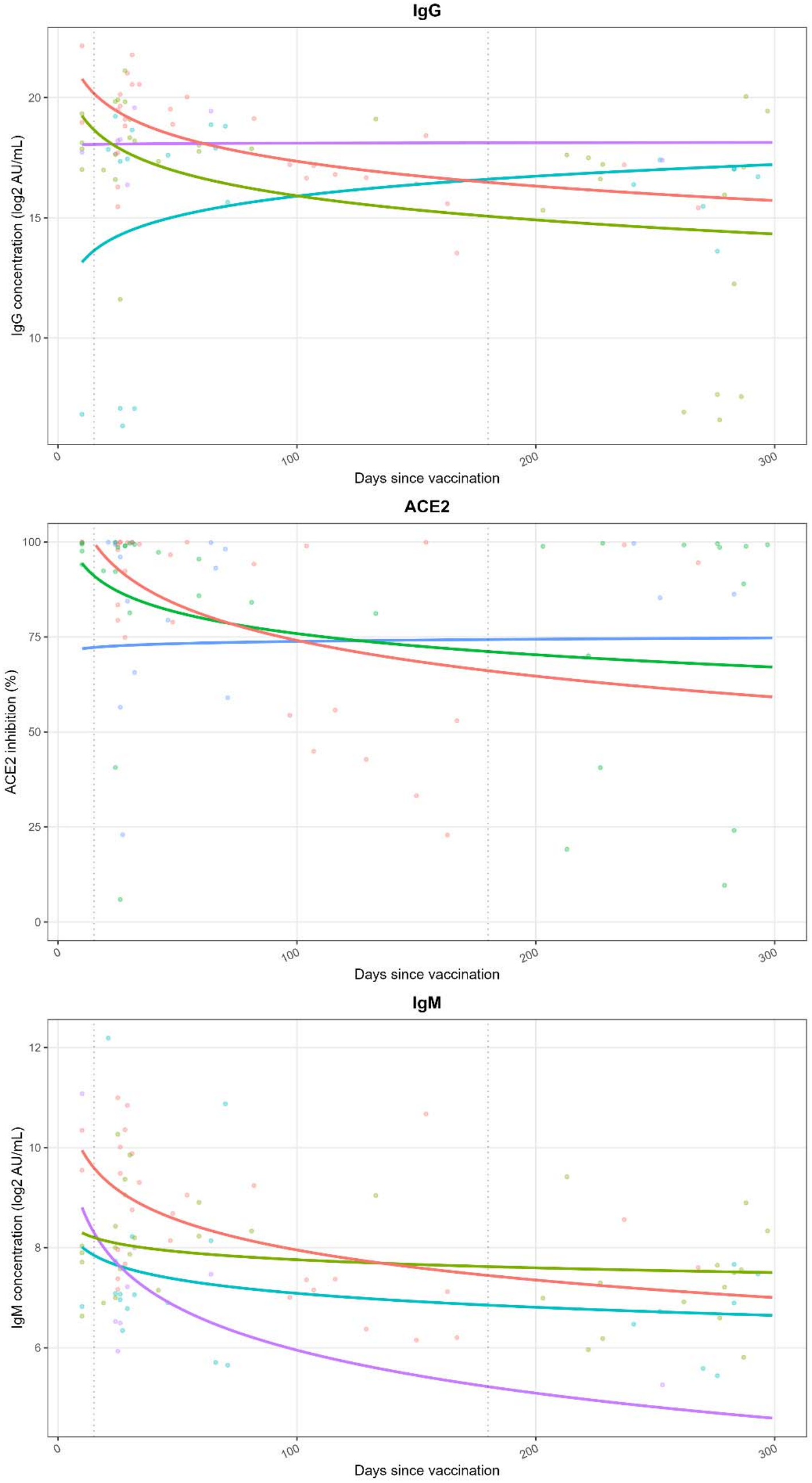
Durability of antibody responses following COVID-19 booster vaccination by vaccine formulation. Observed antibody measurements and fitted power-law regression models describing changes in (A) IgG, (B) IgM, and (C) ACE2 percent inhibition over time following vaccination with the monovalent, bivalent, 2023–2024, and 2024–2025 booster formulations. Lines represent model-predicted trajectories, and points represent observed antibody measurements. Serum samples collected 10–299 days after vaccination were included if obtained before any subsequent vaccination or SARS-CoV-2 infection. The fitted ACE2 trajectory for the 2024–2025 formulation was not displayed because of insufficient data.

Longitudinal power-law models demonstrated variation in antibody waning across vaccine formulations (Figure 5, Table S9). For IgG, following original monovalent vaccination, predicted antibody concentrations decreased by approximately 93% at day 180 and 96% at day 300 relative to peak post-vaccination level (β = −1.03; 95% CI: −1.51 to −0.55; P = 0.024). Similar reductions were estimated following bivalent vaccination (approximately 92% and 95% at days 180 and 300, respectively), with no significant difference in the waning trajectory compared with monovalent vaccination. Following the 2023–24 and 2024–25 formulation, predicted IgG concentrations remained relatively stable. For ACE2 inhibition, significant waning was observed following monovalent vaccination (β = −9.40; 95% CI: −18.0 to −0.8; P = 0.032). The predicted reduction relative to peak titer was approximately 34% at day 180 and 41% at day 300. Following bivalent vaccination, ACE2 inhibition showed predicted decreases of approximately 20% and 24% at days 180 and 300. For the 2023–24 formulation, ACE2 inhibition remained relatively stable. IgM concentrations declined significantly following monovalent vaccination (β= −0.6; 95% CI: −1.0 to −0.2; P = 0.002), corresponding to predicted reductions of approximately 78% and 83% at days 180 and 300 relative to peak titer. The estimated declines were smaller following bivalent vaccination and the 2023–24. Following the 2024–25 formulation, predicted IgM reductions were approximately 89.3% and 93.3% at days 180 and 300, respectively.

Antibody landscapes for individual SARS-CoV-2 variants showed broadly similar patterns of IgG changes across strains within each vaccine formulation, although the magnitude of change varied (Figure 6, Table S10). Following monovalent vaccination, IgG fold changes from peak titer to later post-vaccination serum ranged from 0.38 to 0.73 across evaluated variants, with no clear evidence of difference in durability against the ancestral strain compared with Omicron variants. Larger declines were observed following bivalent vaccination, with fold changes of 0.04–0.05 for most variants, including both vaccine-related and antigenically distant strains. Late IgG concentrations were generally higher following the 2023–24 formulation, while relatively modest declines were observed following the 2024–25 formulation (fold changes, 0.66–0.88).

**Figure 6.**
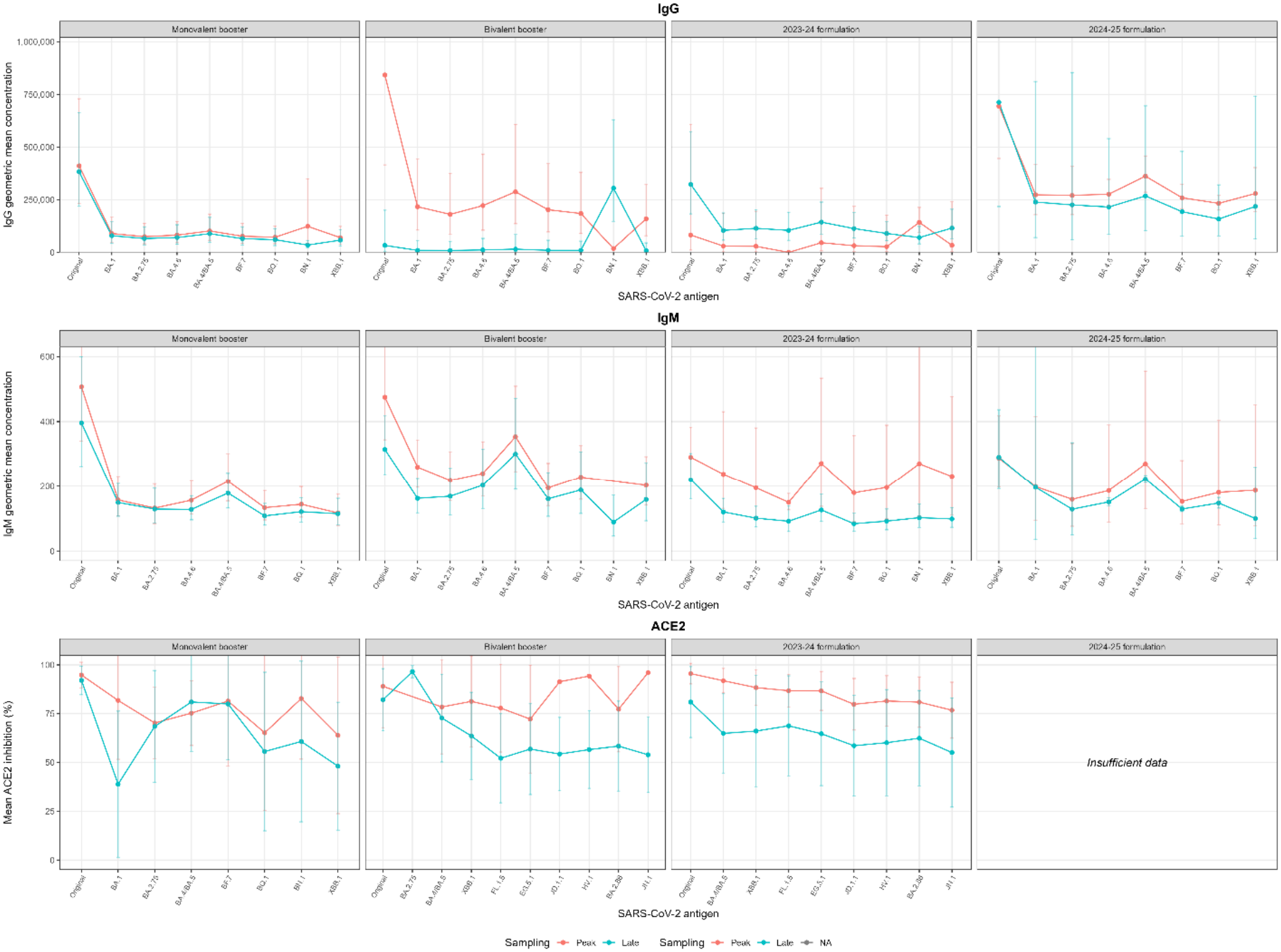
Changes in antibody responses from peak to later time points following COVID-19 booster vaccination across SARS-CoV-2 variants. Antibody responses at peak post-vaccination (Peak) and later post-vaccination (Late) time points are shown for (A) IgG, (B) IgM, and (C) ACE2 percent inhibition across SARS-CoV-2 variants. Panels are presented separately for the original monovalent, bivalent, 2023–2024, and 2024–2025 vaccine formulations. Peak samples were selected as those collected closest to day 15 within 0–100 days after vaccination. Late samples were selected as the latest available eligible samples following vaccination, before any subsequent vaccination or SARS-CoV-2 infection. Lines represent geometric mean concentrations for IgG and IgM or mean percent inhibition for ACE2. ACE2 responses for the 2024–2025 formulation were not displayed because of insufficient data.

ACE2 inhibition landscapes demonstrated greater strain-specific variability, particularly between the ancestral strain, vaccine-related variants, and antigenically distant strains (Figure 6, Table S10). Following monovalent and bivalent vaccination, changes in ACE2 inhibition varied across evaluated variants without a consistent pattern of decline. Following the 2023–24 formulation, ACE2 inhibition generally declined across variants, with mean changes ranging from −11.4% to −27.2 %, suggesting that functional antibody activity decreased against both earlier and contemporary variants rather than exclusively against antigenically distant strains. Limited measurements following the 2024–25 formulation prevented reliable comparisons across strains.

## Discussion

In this longitudinal study of highly vaccinated adults receiving up to seven COVID-19 vaccine doses over five years, we found that repeated booster vaccination maintained high binding and functional antibody responses over time. Although post-vaccination antibody fold-rise, while present, decreased as pre-vaccination antibody concentrations increased, no evidence of reduced immunogenicity was observed after adjusting for baseline antibody levels. Repeated vaccination also generated cross-reactive antibody responses against antigenically distinct SARS-CoV-2 variants, consistent with backboosting, while both IgG binding antibodies and ACE2 inhibition remained durable over time, supporting durable antibody responses following repeated booster vaccination. Our study suggests that repeated vaccination continues to reinforce humoral immunity despite increasingly complex immune histories shaped by prior vaccination and infection.

A key finding of this study that participants with higher baseline IgG concentrations generally experienced smaller antibody increases following booster vaccination, particularly after earlier booster doses. Most previous COVID-19 immunogenicity studies have compared post-vaccination antibody titers or fold-rise without explicitly accounting for baseline immunity, despite evidence that pre-existing immunity strongly influences the magnitude of post-vaccination responses^23,24^. To address this limitation, we adjusted vaccine responses for pre-vaccination antibody concentrations, allowing responses across successive vaccine doses to be compared while accounting for differences in baseline immunity. After adjusting for pre-vaccination antibody concentrations, we did not observe statistically significant changes in immunogenicity across increasing vaccine doses. These findings suggest that the smaller observed antibody increases following repeated vaccination likely reflects a vaccine ceiling effect, whereby individuals with high pre-existing antibody levels have less capacity for further increases following vaccination, rather than an inhibition of immune responses. Similar observations have been reported following repeated COVID-19 booster vaccination, where pre-existing immunity strongly influences post-vaccination antibody responses following bivalent and updated vaccine formulations^23–25^.

Repeated vaccination also generated broad cross-reactive antibody responses across antigenically distinct SARS-CoV-2 variants. Increases in antibody responses were observed not only against vaccine-related variants but also against antigenically distinct variants, particularly the original strain, despite these variants not being included in current vaccine formulations. This pattern is consistent with backboosting, whereby vaccination recalls antibodies generated through previous SARS-CoV-2 infections or vaccinations against related viral strains^10^. Similar immune imprinting has been described following repeated SARS-CoV-2 vaccination, where booster vaccination preferentially recalls antibodies directed toward previously encountered spike epitopes while maintaining responses against updated vaccine antigens^26–28^. The increasing proportion of participants with prior SARS-CoV-2 infection over the study period also reflects the increasingly complex immune histories now present in highly vaccinated populations. Previous studies have shown that hybrid immunity generated through combinations of vaccination and infection is associated with broader and more durable antibody responses^5,28–30^.

An important observation was that antibody responses remained durable following repeated vaccination. Although IgG concentrations and ACE2 inhibition declined over time following peak post-vaccination responses, antibodies remained durable across most participants and evaluated variants throughout follow-up. Functional antibody activity measured by ACE2 inhibition demonstrated similar patterns, suggesting that repeated booster vaccination supports sustained functional as well as s antibody responses. These findings are consistent with previous studies demonstrating an initial decline in SARS-CoV-2 antibody concentrations post vaccination or infection followed by stabilization over time^5,8,31–33^. Our findings extend these observations by demonstrating durable antibody responses following repeated annual booster vaccination across multiple sequential vaccine doses.

This study evaluated repeated COVID-19 vaccination within a cohort of individuals receiving up to seven vaccine doses with longitudinal pre- and post-vaccination serum specimens collected over five years. Most previous COVID-19 immunogenicity studies have focused on responses following one or two updated booster doses^23–25^, whereas relatively few studies have examined antibody responses following repeated annual vaccination across multiple years. The availability of serial serum samples, detailed vaccination histories, documented SARS-CoV-2 infections, and paired measurements of both binding antibodies and functional ACE2 inhibition allowed us to characterize immunogenicity, cross-reactivity, and antibody durability within the same individuals across multiple booster doses. As annual COVID-19 vaccination becomes increasingly common, longitudinal studies of repeatedly vaccinated populations will become increasingly important for understanding long-term immune responses and informing future vaccine strategies.

In conclusion, repeated COVID-19 booster vaccination maintained stable baseline-adjusted immunogenicity while supporting broad cross-reactive and durable antibody responses in a highly vaccinated population. These findings suggest that smaller post-vaccination antibody increases largely reflect higher baseline immunity rather than diminished vaccine responsiveness. As annual COVID-19 vaccination continues, accounting for pre-vaccination immunity will be essential for accurately evaluating vaccine-induced antibody responses and understanding the long-term effects of repeated booster vaccination on population immunity.

## Author Contributions

Conceptualization: Y.Y., E.T.M.

Lab testing: C.J.

Project Administration: A.C.

Data Curation: A.C., M.S., E.T.G., Y.Y.

Analysis: Y.Y.

Visualization: Y.Y.

Original draft writing: Y.Y.

Review and editing: E.T.H., C.J., A.C., M.S., E.T.G., A.S.M.

Funding acquisition: E.T.M., A.S.M.

Supervision: E.T.M.

## Ethical statement

The University of Michigan Institutional Review Board gave ethical approval for this work. Informed consent was obtained at initial enrollment. Parents and legal guardians provided written informed consent for both themselves and their dependents. Children ages 7-9 provided consent verbally, while those 10-13 signed a separate, age-appropriate assent document. Adolescents ages 14-17 read and signed the full informed consent document.

## Finding statement

This study was supported by funding from the National Center for Immunisation and Respiratory Diseases, US Centers for Disease Control and Prevention (75D30122C13149), and the National Institute of Allergy and Infectious Diseases and the National Institutes of Health (75N93021C00015).

## Data availability

The data used in this study can be made available upon request. Due to Institutional Review Board (IRB) regulations, data access is controlled. Per the guidelines of the Centers for Excellence in Influenza Research and Response (CEIRR) Network, individuals seeking access must complete a data and specimen collaboration form. Requests received will be reviewed by the study investigators. For enquiries, please contact. Requests will generally receive an initial response within 3–5 business days.

## Acknowledgements

We would like to thank all individuals for their participation in the HIVE study.

## Reference

1. ACIP COVID-19 Vaccine Recommendations | CDC. May 21, 2024. Accessed July 22, 2024. https://www.cdc.gov/vaccines/hcp/acip-recs/vacc-specific/covid-19.html

2. Research C for BE and. COVID-19 Vaccines (2025-2026 Formula) for Use in the United States Beginning in Fall 2025. FDA. Published online May 22, 2025. Accessed August 12, 2025. https://www.fda.gov/vaccines-blood-biologics/industry-biologics/covid-19-vaccines-2025-2026-formula-use-united-states-beginning-fall-2025

3. Research C for BE and. COVID-19 Vaccines for 2024-2025. FDA. Published online February 5, 2025. Accessed August 12, 2025. https://www.fda.gov/emergency-preparedness-and-response/coronavirus-disease-2019-covid-19/covid-19-vaccines-2024-2025

4. Commissioner O of the. Coronavirus (COVID-19) Update: FDA Authorizes Changes to Simplify Use of Bivalent mRNA COVID-19 Vaccines. FDA. April 18, 2023. Accessed July 5, 2024. https://www.fda.gov/news-events/press-announcements/coronavirus-covid-19-update-fda-authorizes-changes-simplify-use-bivalent-mrna-covid-19-vaccines

5. Moore M, Anderson L, Schiffer JT, Matrajt L, Dimitrov D. Durability of COVID-19 vaccine and infection induced immunity: A systematic review and meta-regression analysis. Vaccine. 2025;54:126966. doi:10.1016/j.vaccine.2025.126966

6. Yang Y, Callear A, Smith M, et al. Longitudinal antibody correlates of SARS-CoV-2 infection in a US household cohort during Omicron waves. medRxiv. Preprint posted online September 11, 2026:2026.09.10.26362752. doi:10.64898/2026.09.10.26362752

7. Yang Y, Juntila C, Callear A, et al. Evaluation of pre-Omicron population immunity for SARS-CoV-2 in the Household Influenza Vaccine Evaluation (HIVE) cohort. medRxiv. Preprint posted online March 11, 2026:2026.03.10.26348071. doi:10.64898/2026.03.10.26348071

8. Yang Y, Risukhina A, Callear A, et al. Antibody boosting and short-term durability following SARS-CoV-2 infections in United States longitudinal cohorts, 2020-2024. medRxiv. Preprint posted online September 20, 2026:2026.09.17.26363304. doi:10.64898/2026.09.17.26363304

9. Thompson MG, Cowling BJ. How repeated influenza vaccination effects might apply to COVID-19 vaccines. The Lancet Respiratory Medicine. 2022;10(7):636–638. doi:10.1016/S2213-2600(22)00162-X

10. Monto AS, Malosh RE, Petrie JG, Martin ET. The Doctrine of Original Antigenic Sin: Separating Good From Evil. J Infect Dis. 2017;215(12):1782–1788. doi:10.1093/infdis/jix173

11. Rössler A, Netzl A, Lasrado N, et al. Nonhuman primate antigenic cartography of SARS-CoV-2. Cell Reports. 2025;44(1). doi:10.1016/j.celrep.2024.115140

12. Rössler A, Netzl A, Knabl L, et al. Characterizing SARS-CoV-2 neutralization profiles after bivalent boosting using antigenic cartography. Nat Commun. 2023;14(1):5224. doi:10.1038/s41467-023-41049-4

13. Nuñez IA, Carlock MA, Allen JD, et al. Impact of age and pre-existing influenza immune responses in humans receiving split inactivated influenza vaccine on the induction of the breadth of antibodies to influenza A strains. PLoS One. 2017;12(11):e0185666. doi:10.1371/journal.pone.0185666

14. Naranbhai V, Garcia-Beltran WF, Chang CC, et al. Comparative Immunogenicity and Effectiveness of mRNA-1273, BNT162b2, and Ad26.COV2.S COVID-19 Vaccines. The Journal of Infectious Diseases. 2022;225(7):1141. doi:10.1093/infdis/jiab593

15. Link-Gelles R. Effectiveness of Monovalent and Bivalent mRNA Vaccines in Preventing COVID-19–Associated Emergency Department and Urgent Care Encounters Among Children Aged 6 Months–5 Years — VISION Network, United States, July 2022–June 2023. MMWR Morb Mortal Wkly Rep. 2023;72. doi:10.15585/mmwr.mm7233a2

16. Plumb ID, Briggs Hagen M, Wiegand R, et al. Effectiveness of a bivalent mRNA vaccine dose against symptomatic SARS-CoV-2 infection among U.S. Healthcare personnel, September 2022–May 2023. Vaccine. Published online November 14, 2023. doi:10.1016/j.vaccine.2023.10.072

17. Rudolph AE, Khan FL, Shah A, et al. Effectiveness of BNT162b2 BA.4/5 Bivalent mRNA Vaccine Against Symptomatic COVID-19 Among Immunocompetent Individuals Testing at a Large US Retail Pharmacy. The Journal of Infectious Diseases. 2024;229(3):648–659. doi:10.1093/infdis/jiad474

18. Monto AS, Foster-Tucker JE, Callear AP, et al. Respiratory Viral Infections From 2015 to 2022 in the HIVE Cohort of American Households: Incidence, Illness Characteristics, and Seasonality. The Journal of Infectious Diseases. Published online August 24, 2024:jiae423. doi:10.1093/infdis/jiae423

19. V-PLEX SARS-CoV-2 Panel 32 (IgG) Kit | Meso Scale Discovery. Accessed August 5, 2025. https://www.mesoscale.com/products/v-plex-sars-cov-2-panel-32-igg-kit-k15668u/

20. V-PLEX SARS-CoV-2 Panel 30 (IgG) Kit | Meso Scale Discovery. Accessed March 16, 2025. https://www.mesoscale.com/products/v-plex-sars-cov-2-panel-30-igg-kit-k15632u/

21. V-PLEX SARS-CoV-2 Panel 38 (IgG) Kit | Meso Scale Discovery. Accessed October 9, 2024. https://www.mesoscale.com/en/products/v-plex-sars-cov-2-panel-38-igg-kit-k15725u/

22. MSD Product Inserts | Meso Scale Discovery. Accessed October 17, 2024. https://www.mesoscale.com/en/support/product_information/product_inserts

23. Lasrado N, Rössler A, McConnell I, et al. Immunogenicity of JN.1 and KP.2 COVID-19 mRNA vaccines against emerging SARS-CoV-2 variants. Vaccine. 2026;69:127997. doi:10.1016/j.vaccine.2025.127997

24. Wang Q, Bowen A, Valdez R, et al. Antibody Response to Omicron BA.4–BA.5 Bivalent Booster. New England Journal of Medicine. 2023;388(6):567–569. doi:10.1056/NEJMc2213907

25. Wrynla XH, Bates TA, Trank-Greene M, et al. Immune imprinting and vaccine interval determine antibody responses to monovalent XBB.1.5 COVID-19 vaccination. Commun Med (Lond). 2025;5(1):182. doi:10.1038/s43856-025-00898-4

26. Röltgen K, Nielsen SCA, Silva O, et al. Immune imprinting, breadth of variant recognition, and germinal center response in human SARS-CoV-2 infection and vaccination. Cell. 2022;185(6):1025–1040.e14. doi:10.1016/j.cell.2022.01.018

27. Wheatley AK, Fox A, Tan HX, et al. Immune imprinting and SARS-CoV-2 vaccine design. Trends Immunol. 2021;42(11):956–959. doi:10.1016/j.it.2021.09.001

28. Voss WN, Mallory MA, Byrne PO, et al. Hybrid immunity to SARS-CoV-2 arises from serological recall of IgG antibodies distinctly imprinted by infection or vaccination. Cell Rep Med. 2024;5(8):101668. doi:10.1016/j.xcrm.2024.101668

29. Reynolds CJ, Pade C, Gibbons JM, et al. Prior SARS-CoV-2 infection rescues B and T cell responses to variants after first vaccine dose. Science. 2021;372(6549):1418–1423. doi:10.1126/science.abh1282

30. Pušnik J, Monzon-Posadas WO, Zorn J, et al. SARS-CoV-2 humoral and cellular immunity following different combinations of vaccination and breakthrough infection. Nat Commun. 2023;14(1):572. doi:10.1038/s41467-023-36250-4

31. Zheng Y, Pan J, Jin M, et al. Efficacy of the neutralizing antibodies after the booster dose on SARS-CoV-2 Omicron variant and a two-year longitudinal antibody study on Wild Type convalescents. Int Immunopharmacol. 2023;119:110151. doi:10.1016/j.intimp.2023.110151

32. Srivastava K, Carreño JM, Gleason C, et al. SARS-CoV-2-infection- and vaccine-induced antibody responses are long lasting with an initial waning phase followed by a stabilization phase. Immunity. 2024;57(3):587–599.e4. doi:10.1016/j.immuni.2024.01.017

33. Russell TW, Townsley H, Hellewell J, et al. Real-time estimation of immunological responses against emerging SARS-CoV-2 variants in the UK: a mathematical modelling study. The Lancet Infectious Diseases. 2025;25(1):80–93. doi:10.1016/S1473-3099(24)00484-5

