## Supplemental materials for "SARS-CoV-2 antibody landscape following multiple vaccinations and infections during Omicron period"

Affiliation:

Supplementary Methods

Vaccine formulation classification

Four formulation groups were defined: original monovalent, bivalent, 2023–24 updated, and 2024–25 updated vaccines. Formulation classification used documented product or formulation information when available, supplemented by vaccination date. The corresponding date periods were September 22, 2021–August 30, 2022; August 31, 2022–September 10, 2023; September 11, 2023–August 21, 2024; and August 22, 2024 onward, respectively. For descriptive analyses, formulation was treated as categorical. For the immunogenicity score analysis, formulations were coded numerically: original monovalent = 1, bivalent = 2, 2023–24 = 3, and 2024–25 = 4.

Antigen selection and antibody measures

Antibody responses were evaluated using antigens corresponding to the SARS-CoV-2 strains included in each COVID-19 vaccine formulation (Table S1). For vaccination events involving the original monovalent formulation, antibody measurements against the ancestral SARS-CoV-2 strain were selected. For bivalent vaccination events, measurements against the ancestral and BA.4/BA.5 strains were considered. For the 2023–24 updated formulation, XBB.1.5-related antigen measurements were selected, while JN.1-related antigen measurements were used for the 2024–25 updated formulation. The formulation-specific antigen selection approach was applied to antibody immunogenicity and durability analyses. IgG and IgM antibody concentrations and ACE2 percent inhibition were evaluated using the corresponding antigen measurements. Antigen-specific analyses examining the breadth of antibody responses across multiple SARS-CoV-2 variants were conducted using all the antigens included in the multiplex assay panels (Table S2).

Selection of pre- and post-vaccination serum samples

Vaccination events were identified by participant identifier and vaccination date. Serum samples were linked to events according to collection dates relative to vaccination. For the peak immunogenicity analysis, post-vaccination samples were collected 10–45 days after vaccination, and the sample closest to day 15 was selected if multiple samples were available. Pre-vaccination samples were selected from eligible specimens collected before the vaccination event.

For antigen-specific analyses, pre-vaccination eligibility incorporated a minimum interval of 90 days after the previous vaccination where applicable. Post-vaccination antigen-specific samples were considered within 180 days after vaccination and before the next documented vaccination. Paired analyses required eligible pre- and post-vaccination measurements for the same vaccination event and antigen.

Descriptive statistics of antibody boosting

IgG and IgM concentrations were summarized as geometric mean concentrations (GMCs), with 95% confidence intervals calculated on the log-scale. For paired observations, the fold change was defined as:

*FCᵢ = Aᵢ,post / Aᵢ,pre*

The geometric mean fold-rise (GMFR) was calculated as:

*GMFR = exp[(1/n) Σᵢ ln(FCᵢ)]*

For ACE2 inhibition, absolute paired change was calculated in percentage points:

*ΔACE2ᵢ = ACE2ᵢ,post − ACE2ᵢ,pre*

Baseline antibody levels and boosting responses

Linear regression evaluated the association between pre-vaccination antibody measurements and subsequent boosting. IgG and IgM pre-vaccination concentrations were log₂-transformed; ACE2 pre-vaccination percent inhibition was retained on its original scale. Models were fitted separately for IgG, IgM, and ACE2 inhibition:

*Yᵢ = α + βXᵢ,pre + εᵢ*

Here Yᵢ is the boosting response, Xᵢ,pre is the baseline antibody measurement, β is the baseline-response slope, and εᵢ is residual error. For IgG and IgM, boosting was expressed on the fold-change scale specified by the analysis; for ACE2, it was expressed as an absolute percentage-point change. Regression coefficients, 95% confidence intervals, and P values were reported.

Baseline-adjusted immunogenicity scores

A two-stage residual-based approach assessed trends in boosting after accounting for baseline antibody levels. First, a linear model predicted the boosting response from baseline antibody measurements. The difference between observed and predicted responses was the immunogenicity score:

*Rᵢ = Yᵢ − Ŷᵢ*

Second, the residual score was regressed on numerically coded vaccine formulation:

*Rᵢ = γ₀ + γ₁Fᵢ + ηᵢ*

Fᵢ was coded from 1 to 4, and γ₁ estimated the average change in residual score per one-unit increase in formulation number. Models were fitted separately for IgG, IgM, and ACE2. Ordinary linear regression was used without participant-level random intercepts or cluster-based variance estimation. The coefficient reflects an ordinal linear trend, not separate pairwise formulation comparisons.

Descriptive antibody landscapes

Descriptive landscapes compared antigen-specific IgG and IgM GMCs and mean ACE2 percent inhibition across SARS-CoV-2 variants and vaccine formulations. Antigens were ordered chronologically by emergence. Separate landscapes compared pre- versus post-vaccination responses and peak versus later follow-up measurements. Where observations were unmatched, differences were interpreted descriptively.

Antigenic cartography-based landscapes

A two-dimensional antigenic map was reconstructed using published neutralization data from single-exposure human sera and live-virus isolates reported by Rössler et al (Figure S1). Neutralization titers were transformed into log₂ fold-drops relative to the maximum titer for each serum sample. Euclidean antigenic coordinates were optimized to represent observed relationships among variants. These coordinates served as a fixed reference for projecting log-transformed IgG concentrations and constructing continuous antibody response surfaces across antigenic space. Landscape height represented estimated antibody concentration.

Peak and later follow-up antibody responses

Durability analyses included eligible serum measurements collected approximately 10–300 days after vaccination. Peak responses were selected from the early post-vaccination window, closest to day 15. The pre-next-vaccination measurement was the latest eligible sample before the subsequent vaccination and at least 60 days after the peak sample. IgG and IgM were summarized as GMCs and fold changes, and ACE2 as mean inhibition and absolute percentage-point differences. Matched peak-to-pre-next comparisons were distinguished from unpaired peak-versus-late summaries.

Power-law waning models

Power-law regression characterized antibody measurements over time since vaccination. Independent regression models were fitted for each vaccine formulation and antibody target. The general model was:

*Yᵢf = β₀f + β₁f log₂(tᵢf) + εᵢf*

Here, Yᵢf represents log₂-transformed IgG or IgM concentrations, or untransformed ACE2 percent inhibition, for observation i in vaccine formulation f; tᵢf represents days since vaccination; β₀f is the formulation-specific intercept; and β₁f is the formulation-specific time slope. For IgG and IgM, the slope represents the change in log₂ antibody concentration per doubling of time since vaccination. For ACE2, the slope represents the percentage-point change in inhibition per doubling of time. Models were fitted independently for the monovalent, bivalent, 2023–24, and 2024–25 vaccine formulations.

Predicted IgG and IgM concentrations were back-transformed to the original scale, while ACE2 predictions remained on the percentage inhibition scale. Predicted responses and 95% confidence intervals were calculated at 15, 180, and 300 days post-vaccination. Relative changes in IgG and IgM were calculated using ratios of model-predicted concentrations, whereas ACE2 changes were expressed as absolute percentage-point differences. Follow-up was restricted to the specified post-vaccination window and censored at subsequent vaccination or documented infection where applicable. Models did not include participant-level random effects.

Sensitivity analyses and model assessment

Leave-one-participant-out cross-validation assessed the influence of individual participants on immunogenicity trends. In each iteration, all observations for one participant were excluded and the immunogenicity model refitted; estimated formulation effects were compared with the full-sample estimate.

Table S1: COVID-19 vaccine dose history and corresponding formulations

| **Dose number^1^** | **Role in vaccination series** | **Vaccine formulation** | **SARS-CoV-2 antigen(s)** | **First FDA authorization/approval** | **Target antigen for immunogenicity analysis** | **Target antigen selected for durability analysis** |
| --- | --- | --- | --- | --- | --- | --- |
| 1 | Primary series, dose 1 | Original monovalent mRNA | Ancestral | December 2020 | Ancestral | Ancestral |
| 2 | Primary series, dose 2 | Original monovalent mRNA | Ancestral | December 2020 | Ancestral | Ancestral |
| 3 | First booster | Original monovalent mRNA | Ancestral | September 2021 | Ancestral | Ancestral |
| 4 | Second booster | Original monovalent mRNA | Ancestral | March 2022 | Ancestral | Ancestral |
| 5 | Third booster | Bivalent mRNA | Ancestral + BA.4/BA.5 | August 31, 2022 | BA.5 | BA.4/BA.5 |
| 6 | Fourth booster | Updated monovalent mRNA | XBB.1.5 | September 11, 2023 | XBB.1.5 | XBB.1.5 |
| 7 | Fifth booster | Updated monovalent mRNA | JN.1/KP.2 lineage | August 22, 2024 | JN.1 | JN.1 |

^1^ Dose number represents the sequential number of COVID-19 vaccine doses received by each participant. Doses 1–2 generally represented the original monovalent primary series; doses 3–4 the original monovalent booster doses; dose 5 the Original/Omicron BA.4/BA.5 bivalent booster; dose 6 the updated XBB.1.5 monovalent vaccine; and dose 7 the updated 2024–2025 JN.1-lineage vaccine. Vaccine formulation at a given dose number could vary according to calendar time and individual vaccination history.

Table S2: Serological sample testing assay summary

| V-PLEX Serology and Neutralization Panels | Assay | Target | Time period |
| --- | --- | --- | --- |
| Panel 32 | IgG, IgM | SARS-CoV-2 Spike  SARS-CoV-2 Spike (B.1.1.529; BA.1; BA.1.15)  SARS-CoV-2 Spike (BA.2.75)  SARS-CoV-2 Spike (BA.2.75.2)  SARS-CoV-2 Spike (BA.4.6)  SARS-CoV-2 Spike (BA.5)  SARS-CoV-2 Spike (BF.7)  SARS-CoV-2 Spike (BQ.1)  SARS-CoV-2 Spike (BQ.1.1)  SARS-CoV-2 Spike (XBB.1) | August 01, 2020 to August 01, 2025 |
| Panel 33 | ACE2 | SARS-CoV-2 S1 RBD  SARS-CoV-2 S1 RBD (B.1.1.529; BA.1; BA.1.15)  SARS-CoV-2 S1 RBD (BA.2.75)  SARS-CoV-2 S1 RBD (BA.2.75.2)  SARS-CoV-2 S1 RBD (BA.4; BA.5)  SARS-CoV-2 S1 RBD (BA.4.6; BF.7)  SARS-CoV-2 S1 RBD (BQ.1)  SARS-CoV-2 S1 RBD (BQ.1.1)  SARS-CoV-2 S1 RBD (XBB.1) | August 01, 2020 to July 21, 2023 |
| Panel 38 | ACE2 | SARS-CoV-2 Spike  SARS-CoV-2 Spike (BA.2.86)  SARS-CoV-2 Spike (BA.5)  SARS-CoV-2 Spike (EG.5.1)  SARS-CoV-2 Spike (FL.1.5.1)  SARS-CoV-2 Spike (XBB.1.5)  SARS-CoV-2 Spike (JN.1)  SARS-CoV-2 Spike (JD.1.1)  SARS-CoV-2 Spike (HV.1) | August 01, 2023 to August 01, 2025 |

Table S3. Demographic characteristics of study participants

| **Characteristic** | **N = 21**^1^ |
| --- | --- |
| **Age, years** | 51.1 (49.8, 55.9) |
| **Gender** |  |
| F | 13 (62%) |
| M | 8 (38%) |
| **Race/ethnicity** |  |
| Missing or Unknown | 17 (81%) |
| Non-Hispanic Black or African American | 1 (4.8%) |
| Non-Hispanic White | 3 (14%) |
| **Any high-risk condition** |  |
| No | 12 (57%) |
| Yes | 9 (43%) |
| Missing | 0 (0%) |
| **Number of children in household** | 2.0 (2.0, 3.0) |
| ^1^Median (Q1, Q3); n (%) | |

Figure S1: SARS-CoV-2 Antigenic cartography base map. Data used this graph were abstract from Rössler A, Netzl A, Lasrado N, et al. Nonhuman primate antigenic cartography of SARS-CoV-2. *Cell Reports*. 2025;44(1). doi:10.1016/j.celrep.2024.115140.

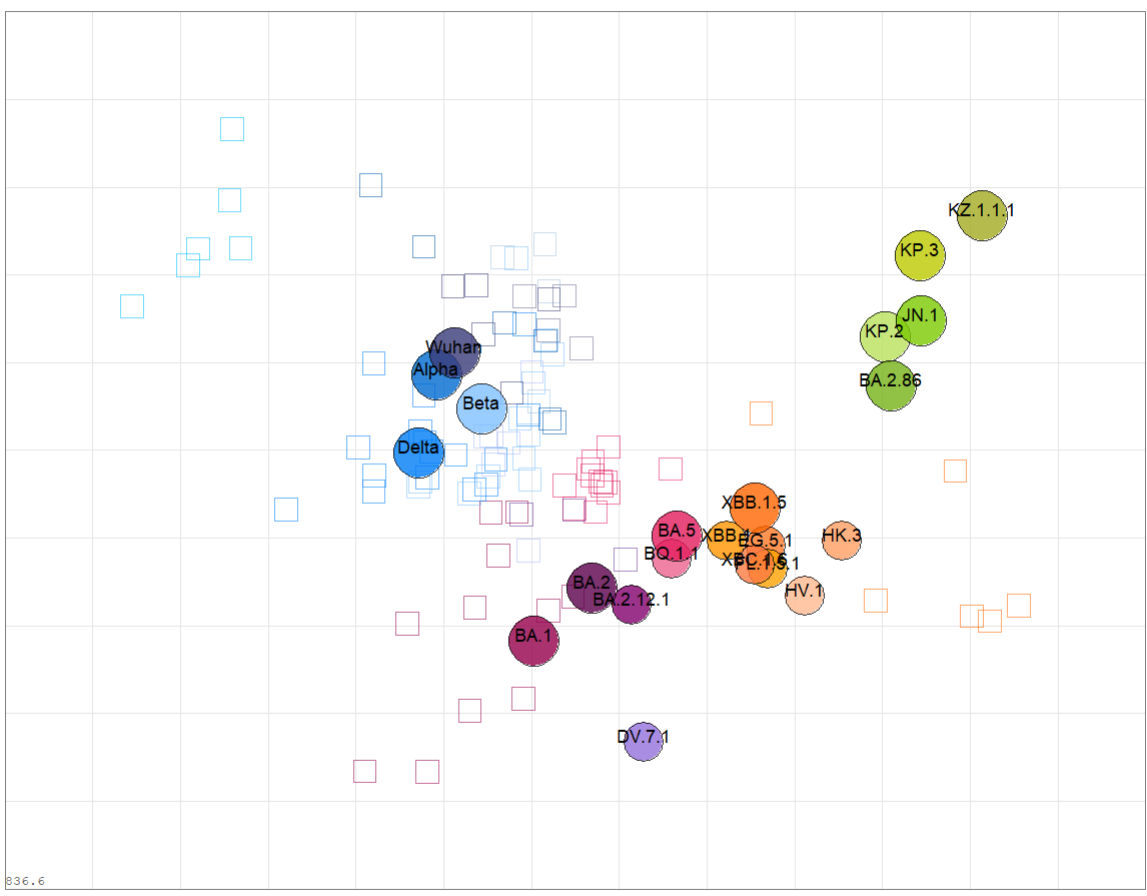

Figure S2: Study participant serological sample collection, vaccination and infection date distribution

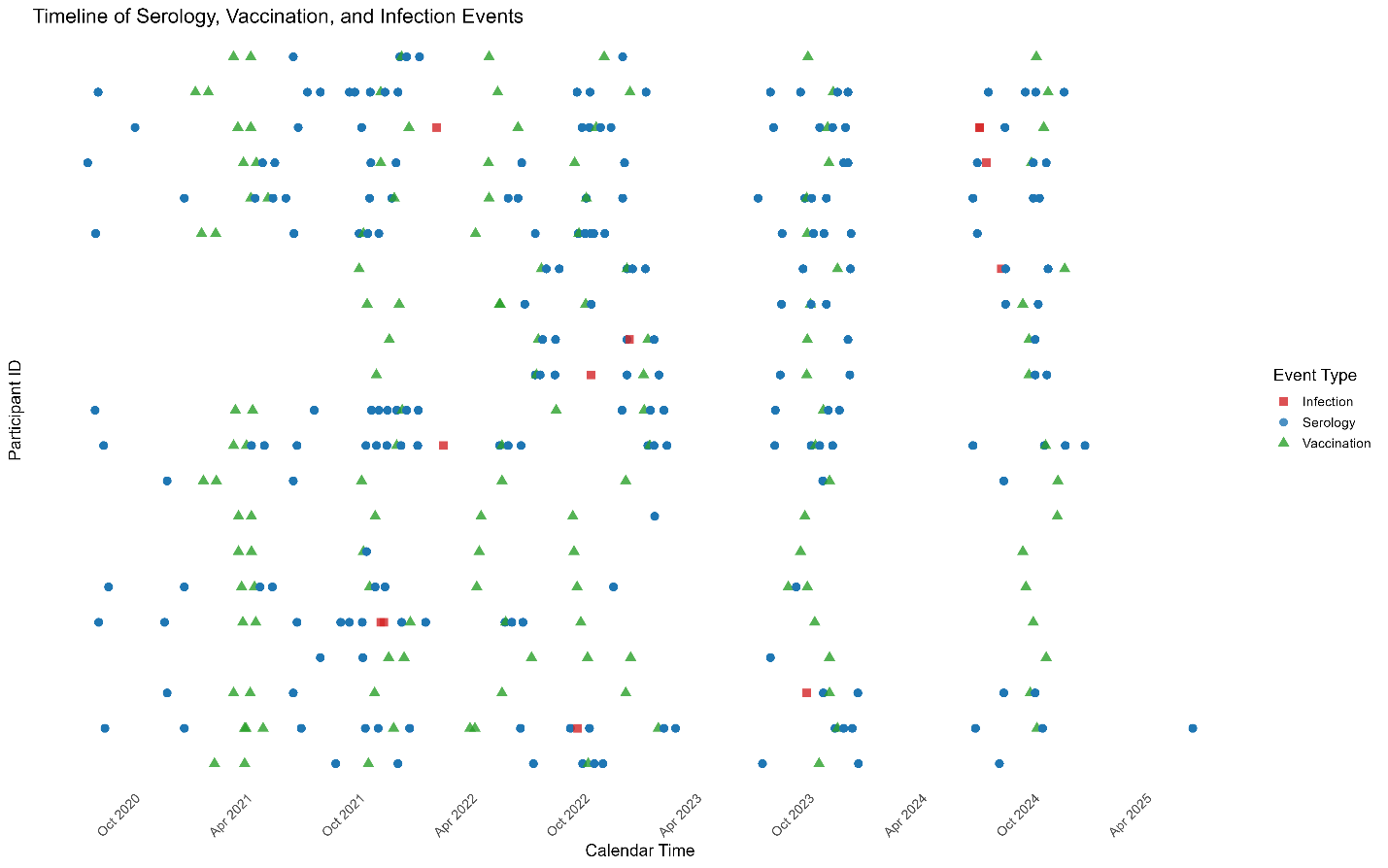

Table S4: Summary statistics of IgG/IgM antibody response among participants by vaccination booster formulation

| Antibody | Vaccine formulation | Vaccination events (n) | Pre-vaccination GMC (95% CI) | Pre-sample days, median (IQR) | Post-vaccination GMC (95% CI) | Post-sample days, median (IQR) | Paired fold-rise or change, median (range) | Prior infection, % |
| --- | --- | --- | --- | --- | --- | --- | --- | --- |
| IgG | Monovalent booster | 19 | 146,009.9 (68,588.7–310,822.3) | 17.0 (8.0–131.0) | 423,569.8 (228,166.8–786,317.1) | 25.0 (7.5–42.5) | 4.5 (0.2–40.3) | 15.8% |
| IgG | Bivalent booster | 14 | 177,159.8 (73,984.8–424,216.8) | 50.5 (15.0–97.8) | 252,231.9 (109,133.7–582,963.3) | 10.0 (9.0–19.0) | 2.0 (0.0–71.4) | 42.9% |
| IgG | 2023-24 formulation | 13 | 3,197.3 (471.0–21,703.7) | 43.0 (13.0–56.0) | 23,458.5 (2,378.8–231,340.6) | 15.5 (8.0–41.0) | 50.6 (0.0–2,659.6) | 23.1% |
| IgG | 2024-25 formulation | 9 | 136,036.2 (102,014.6–181,403.9) | 43.0 (27.0–88.0) | 291,974.4 (191,846.8–444,360.0) | 24.5 (12.8–25.8) | 2.4 (0.8–3.2) | 66.7% |
| IgM | Monovalent booster | 19 | 474.5 (299.4–751.9) | 17.0 (8.0–131.0) | 429.1 (264.8–695.1) | 25.0 (7.5–42.5) | 0.9 (0.3–4.8) | 15.8% |
| IgM | Bivalent booster | 14 | 308.4 (171.7–554.1) | 50.5 (15.0–97.8) | 294.8 (190.5–456.3) | 10.0 (9.0–19.0) | 1.4 (0.3–2.7) | 42.9% |
| IgM | 2023-24 formulation | 13 | 143.4 (95.4–215.6) | 43.0 (13.0–56.0) | 270.3 (119.7–610.2) | 15.5 (8.0–41.0) | 1.7 (0.2–30.0) | 23.1% |
| IgM | 2024-25 formulation | 9 | 161.0 (71.9–360.5) | 43.0 (27.0–88.0) | 125.1 (81.4–192.2) | 24.5 (12.8–25.8) | 1.1 (0.1–2.0) | 66.7% |

| Antibody | Vaccine formulation | Vaccination events (n) | Pre-vaccination ACE2 mean (95% CI) | Pre-sample days, median (IQR) | Post-vaccination ACE2 mean (95% CI) | Post-sample days, median (IQR) | Paired fold-rise or change, median (range) | Prior infection, % |
| --- | --- | --- | --- | --- | --- | --- | --- | --- |
| ACE2 | Monovalent booster | 19 | 81.1 (69.3–93.0) | 17.0 (8.0–131.0) | 91.2 (82.0–100.4) | 25.0 (7.5–42.5) | 2.7 (-49.6–98.6) | 15.8% |
| ACE2 | Bivalent booster | 14 | 71.8 (55.0–88.6) | 50.5 (15.0–97.8) | 81.9 (64.9–98.8) | 10.0 (9.0–19.0) | 0.6 (-74.9–97.4) | 42.9% |
| ACE2 | 2023-24 formulation | 13 | 51.4 (30.8–71.9) | 43.0 (13.0–56.0) | 70.8 (48.1–93.5) | 15.5 (8.0–41.0) | 30.0 (-34.7–99.9) | 23.1% |
| ACE2 | 2024-25 formulation | 9 | 79.7 (60.4–98.9) | 65.5 (31.8–97.0) | 81.0 (63.0–98.9) | 25.0 (8.0–26.0) | 7.3 (-47.5–39.7) | 66.7% |

Table S5: Association between pre-vaccination antibody levels and post-vaccination boosting, by vaccine formulation.

| Vaccine formulation | Antibody | n | Slope | Slope (95% CI) | P value |
| --- | --- | --- | --- | --- | --- |
| Monovalent booster | IgG | 15 | -0.5912 | -0.5912 (-0.9372–-0.2452) | 0.0027 |
| Monovalent booster | IgM | 15 | -0.3441 | -0.3441 (-0.7003–0.0121) | 0.0571 |
| Bivalent booster | IgG | 13 | -0.8131 | -0.8131 (-1.3978–-0.2283) | 0.0108 |
| Bivalent booster | IgM | 13 | -0.4290 | -0.4290 (-0.7450–-0.1130) | 0.0123 |
| 2023-24 formulation | IgG | 10 | -1.1625 | -1.1625 (-1.9842–-0.3408) | 0.0115 |
| 2023-24 formulation | IgM | 10 | -1.1837 | -1.1837 (-2.4189–0.0514) | 0.0581 |
| 2024-25 formulation | IgG | 6 | -0.5446 | -0.5446 (-1.9410–0.8517) | 0.3397 |
| 2024-25 formulation | IgM | 6 | -0.7055 | -0.7055 (-1.1294–-0.2817) | 0.0099 |

| Vaccine formulation | Antibody | n | Slope | Slope (95% CI) | P value |
| --- | --- | --- | --- | --- | --- |
| Monovalent booster | ACE2 | 15 | -1.1503 | -1.1503 (-1.5137–-0.7868) | 0.0000 |
| Bivalent booster | ACE2 | 13 | -0.9161 | -0.9161 (-1.5343–-0.2979) | 0.0076 |
| 2023-24 formulation | ACE2 | 10 | -0.6395 | -0.6395 (-1.4052–0.1262) | 0.0903 |
| 2024-25 formulation | ACE2 | 5 | -0.9852 | -0.9852 (-2.4389–0.4685) | 0.1199 |

Table S6: Immunogenicity score model results by antibody measurement and numerical vaccine formulation

| Antibody | Serum number | Log₂-transformed baseline antibody slope (95% CI)^1^ | LM^2^ P value | Vaccine formulation effect (95% CI) | P value |
| --- | --- | --- | --- | --- | --- |
| IgG | 44 | −0.73 (−0.98, −0.49) | <0.001 | −0.58 (−1.49, 0.34) | 0.208 |
| IgM | 44 | −0.53 (−0.76, −0.29) | <0.001 | −0.20 (−0.56, 0.16) | 0.269 |
| ACE2 | 43 | −0.84 (−1.09, −0.58) | <0.001 | −4.71 (−12.95, 3.53) | 0.255 |

^1^Linear regression

^2^Linear mixed effect model

Table S7: Antigen-specific IgG and IgM antibody concentrations and boosting by vaccine formulation

| Antibody | Vaccine formulation | Antigen | Pre GMC (95% CI) | Post GMC (95% CI) | Paired n; increased n (%) | GMFR (95% CI) |
| --- | --- | --- | --- | --- | --- | --- |
| IgG | Monovalent booster | BA.1 | 14,090.75 (5,190.92–38,249.32) | 90,688.75 (42,090.82–195,397.68) | 12; 11 (91.7%) | 6.44 (3.08–13.45) |
| IgG | Monovalent booster | BA.2.75 | 12,728.59 (4,686.74–34,569.29) | 77,123.84 (36,533.70–162,810.99) | 12; 12 (100.0%) | 6.06 (2.98–12.32) |
| IgG | Monovalent booster | BA.4.6 | 13,940.15 (5,228.59–37,166.36) | 84,326.87 (41,727.66–170,415.04) | 12; 12 (100.0%) | 6.05 (3.02–12.11) |
| IgG | Monovalent booster | BA.4/BA.5 | 18,056.37 (6,763.39–48,205.50) | 105,689.13 (52,489.37–212,808.64) | 12; 12 (100.0%) | 5.85 (2.90–11.82) |
| IgG | Monovalent booster | BF.7 | 13,137.28 (4,956.78–34,818.57) | 79,539.83 (39,405.87–160,549.31) | 12; 12 (100.0%) | 6.05 (3.01–12.18) |
| IgG | Monovalent booster | BN.1 | 59,036.99 | 165,345.56 | 1; 1 (100.0%) | 2.80 |
| IgG | Monovalent booster | BQ.1 | 11,616.97 (4,852.84–27,809.25) | 72,981.71 (37,977.98–140,247.82) | 12; 12 (100.0%) | 6.28 (3.25–12.13) |
| IgG | Monovalent booster | Original | 81,389.98 (29,594.01–223,840.23) | 426,966.73 (218,971.79–832,530.02) | 12; 12 (100.0%) | 5.25 (2.70–10.18) |
| IgG | Monovalent booster | XBB.1 | 11,425.24 (4,407.90–29,614.14) | 69,745.31 (35,293.80–137,826.16) | 12; 12 (100.0%) | 6.10 (3.02–12.34) |
| IgG | Bivalent booster | BA.1 | 94,105.52 (43,616.02–203,041.18) | 228,237.47 (87,757.90–593,591.54) | 11; 10 (90.9%) | 2.43 (0.81–7.25) |
| IgG | Bivalent booster | BA.2.75 | 73,231.99 (32,498.94–165,018.42) | 184,460.19 (69,277.00–491,152.30) | 11; 10 (90.9%) | 2.52 (0.84–7.52) |
| IgG | Bivalent booster | BA.4.6 | 87,678.04 (37,887.30–202,902.77) | 237,064.28 (88,337.56–636,189.97) | 11; 10 (90.9%) | 2.70 (0.95–7.72) |
| IgG | Bivalent booster | BA.4/BA.5 | 114,132.44 (48,140.84–270,585.52) | 304,889.33 (112,614.84–825,446.28) | 11; 10 (90.9%) | 2.67 (0.93–7.63) |
| IgG | Bivalent booster | BF.7 | 78,979.06 (34,379.69–181,435.40) | 215,836.69 (81,481.34–571,731.83) | 11; 10 (90.9%) | 2.73 (0.96–7.74) |
| IgG | Bivalent booster | BN.1 | 16,327.64 | 18,450.69 | 1; 1 (100.0%) | 1.13 |
| IgG | Bivalent booster | BQ.1 | 73,941.40 (32,176.66–169,916.01) | 195,680.37 (74,962.03–510,802.72) | 11; 10 (90.9%) | 2.65 (0.92–7.63) |
| IgG | Bivalent booster | Original | 449,089.21 (224,959.63–896,521.37) | 870,026.32 (334,552.94–2,262,559.08) | 11; 10 (90.9%) | 1.94 (0.74–5.10) |
| IgG | Bivalent booster | XBB.1 | 60,448.82 (27,965.00–130,665.46) | 166,081.62 (65,177.71–423,198.40) | 11; 10 (90.9%) | 2.75 (0.95–7.95) |
| IgG | 2023-24 formulation | BA.1 | 2,054.20 (265.00–15,923.39) | 21,280.53 (2,525.36–179,325.22) | 10; 7 (70.0%) | 10.36 (0.42–257.35) |
| IgG | 2023-24 formulation | BA.2.75 | 1,944.20 (221.03–17,101.36) | 20,018.83 (2,139.55–187,307.78) | 10; 7 (70.0%) | 10.30 (0.38–276.68) |
| IgG | 2023-24 formulation | BA.4.6 | 5,983.90 (63.78–561,378.86) | 150.26 (127.13–177.60) | 3; 1 (33.3%) | 0.03 (0.00–2.00) |
| IgG | 2023-24 formulation | BA.4/BA.5 | 3,661.78 (467.38–28,688.62) | 32,825.97 (3,557.78–302,870.05) | 10; 8 (80.0%) | 8.96 (0.38–212.50) |
| IgG | 2023-24 formulation | BF.7 | 2,202.58 (261.57–18,547.16) | 21,566.97 (2,309.75–201,378.33) | 10; 7 (70.0%) | 9.79 (0.35–271.19) |
| IgG | 2023-24 formulation | BQ.1 | 2,280.32 (287.33–18,097.46) | 18,522.76 (2,074.23–165,407.13) | 10; 7 (70.0%) | 8.12 (0.32–206.71) |
| IgG | 2023-24 formulation | Original | 7,177.59 (781.44–65,926.59) | 58,117.50 (5,561.17–607,362.15) | 10; 8 (80.0%) | 8.10 (0.27–241.55) |
| IgG | 2023-24 formulation | XBB.1 | 1,760.80 (226.70–13,676.44) | 24,102.24 (2,385.83–243,487.15) | 10; 8 (80.0%) | 13.69 (0.51–364.67) |
| IgG | 2024-25 formulation | BA.1 | 120,305.88 (77,510.43–186,729.75) | 283,153.07 (172,712.79–464,213.80) | 6; 6 (100.0%) | 2.35 (1.55–3.56) |
| IgG | 2024-25 formulation | BA.2.75 | 138,261.14 (92,666.31–206,290.11) | 284,130.59 (176,875.93–456,422.72) | 6; 5 (83.3%) | 2.06 (1.30–3.24) |
| IgG | 2024-25 formulation | BA.4.6 | 146,535.01 (91,755.51–234,018.75) | 285,543.58 (220,749.15–369,356.51) | 6; 5 (83.3%) | 1.95 (1.19–3.18) |
| IgG | 2024-25 formulation | BA.4/BA.5 | 188,805.74 (123,523.92–288,588.70) | 374,296.96 (286,321.40–489,304.01) | 6; 5 (83.3%) | 1.98 (1.31–2.99) |
| IgG | 2024-25 formulation | BF.7 | 148,982.80 (96,937.51–228,970.96) | 266,736.65 (206,585.55–344,401.83) | 6; 5 (83.3%) | 1.79 (1.15–2.79) |
| IgG | 2024-25 formulation | BQ.1 | 111,143.13 (72,107.82–171,310.08) | 236,447.22 (198,982.98–280,965.18) | 6; 6 (100.0%) | 2.13 (1.40–3.23) |
| IgG | 2024-25 formulation | Original | 400,676.89 (279,682.62–574,014.83) | 738,363.38 (446,061.40–1,222,209.50) | 6; 5 (83.3%) | 1.84 (1.07–3.17) |
| IgG | 2024-25 formulation | XBB.1 | 146,850.30 (103,753.62–207,848.28) | 291,974.41 (191,846.84–444,359.99) | 6; 6 (100.0%) | 1.99 (1.35–2.93) |

| Antibody | Vaccine formulation | Antigen | Pre GMC (95% CI) | Post GMC (95% CI) | Paired n; increased n (%) | GMFR (95% CI) |
| --- | --- | --- | --- | --- | --- | --- |
| IgM | Monovalent booster | BA.1 | 175.81 (109.23–282.98) | 188.43 (120.87–293.74) | 12; 7 (58.3%) | 1.07 (0.68–1.69) |
| IgM | Monovalent booster | BA.2.75 | 141.75 (80.63–249.18) | 161.72 (95.20–274.73) | 12; 7 (58.3%) | 1.14 (0.64–2.04) |
| IgM | Monovalent booster | BA.4.6 | 153.91 (90.43–261.96) | 165.44 (107.14–255.46) | 12; 6 (50.0%) | 1.07 (0.71–1.62) |
| IgM | Monovalent booster | BA.4/BA.5 | 200.18 (118.66–337.72) | 223.58 (143.02–349.50) | 12; 7 (58.3%) | 1.12 (0.76–1.64) |
| IgM | Monovalent booster | BF.7 | 130.18 (75.33–224.97) | 145.85 (93.80–226.80) | 12; 7 (58.3%) | 1.12 (0.70–1.79) |
| IgM | Monovalent booster | BQ.1 | 144.91 (88.31–237.81) | 152.48 (100.42–231.51) | 12; 6 (50.0%) | 1.05 (0.73–1.52) |
| IgM | Monovalent booster | Original | 541.75 (284.66–1,031.03) | 576.32 (344.99–962.75) | 12; 6 (50.0%) | 1.06 (0.71–1.58) |
| IgM | Monovalent booster | XBB.1 | 142.57 (86.51–234.95) | 140.40 (86.77–227.18) | 12; 7 (58.3%) | 0.98 (0.59–1.65) |
| IgM | Bivalent booster | BA.1 | 160.85 (108.22–239.06) | 283.31 (200.60–400.13) | 11; 11 (100.0%) | 1.76 (1.41–2.20) |
| IgM | Bivalent booster | BA.2.75 | 152.20 (92.27–251.05) | 244.46 (158.12–377.95) | 11; 10 (90.9%) | 1.61 (1.21–2.13) |
| IgM | Bivalent booster | BA.4.6 | 202.62 (96.20–426.78) | 260.37 (168.00–403.53) | 11; 9 (81.8%) | 1.28 (0.83–1.98) |
| IgM | Bivalent booster | BA.4/BA.5 | 282.69 (130.68–611.53) | 375.67 (234.87–600.87) | 11; 9 (81.8%) | 1.33 (0.88–2.01) |
| IgM | Bivalent booster | BF.7 | 162.43 (77.86–338.86) | 215.20 (142.71–324.51) | 11; 9 (81.8%) | 1.32 (0.85–2.06) |
| IgM | Bivalent booster | BQ.1 | 190.32 (84.87–426.79) | 245.70 (154.00–391.99) | 11; 9 (81.8%) | 1.29 (0.81–2.06) |
| IgM | Bivalent booster | Original | 294.10 (204.37–423.23) | 448.08 (318.37–630.62) | 11; 11 (100.0%) | 1.52 (1.36–1.70) |
| IgM | Bivalent booster | XBB.1 | 149.72 (83.45–268.60) | 218.73 (135.68–352.61) | 11; 10 (90.9%) | 1.46 (1.17–1.82) |
| IgM | 2023-24 formulation | BA.1 | 139.85 (95.52–204.74) | 268.36 (140.04–514.26) | 10; 7 (70.0%) | 1.92 (0.85–4.33) |
| IgM | 2023-24 formulation | BA.2.75 | 140.24 (83.37–235.89) | 222.92 (105.55–470.80) | 10; 6 (60.0%) | 1.59 (0.65–3.88) |
| IgM | 2023-24 formulation | BA.4.6 | 134.57 (56.23–322.03) | 150.26 (127.13–177.60) | 3; 2 (66.7%) | 1.12 (0.53–2.36) |
| IgM | 2023-24 formulation | BA.4/BA.5 | 232.66 (133.00–407.00) | 316.18 (147.15–679.38) | 10; 6 (60.0%) | 1.36 (0.62–2.97) |
| IgM | 2023-24 formulation | BF.7 | 128.50 (76.73–215.20) | 209.06 (96.20–454.32) | 10; 7 (70.0%) | 1.63 (0.74–3.59) |
| IgM | 2023-24 formulation | BQ.1 | 142.04 (79.86–252.64) | 223.54 (102.90–485.63) | 10; 7 (70.0%) | 1.57 (0.66–3.76) |
| IgM | 2023-24 formulation | Original | 268.66 (200.14–360.65) | 311.94 (236.77–410.97) | 10; 7 (70.0%) | 1.16 (0.71–1.90) |
| IgM | 2023-24 formulation | XBB.1 | 129.20 (70.77–235.86) | 270.25 (119.70–610.16) | 10; 8 (80.0%) | 2.09 (0.73–5.97) |
| IgM | 2024-25 formulation | BA.1 | 135.70 (76.39–241.08) | 147.47 (87.21–249.37) | 6; 4 (66.7%) | 1.09 (0.68–1.73) |
| IgM | 2024-25 formulation | BA.2.75 | 117.02 (52.31–261.75) | 114.64 (76.25–172.37) | 6; 4 (66.7%) | 0.98 (0.60–1.60) |
| IgM | 2024-25 formulation | BA.4.6 | 141.23 (57.25–348.43) | 137.49 (83.31–226.91) | 6; 4 (66.7%) | 0.97 (0.56–1.70) |
| IgM | 2024-25 formulation | BA.4/BA.5 | 199.63 (83.76–475.81) | 200.65 (120.61–333.79) | 6; 4 (66.7%) | 1.01 (0.61–1.65) |
| IgM | 2024-25 formulation | BF.7 | 118.84 (54.82–257.63) | 118.36 (80.44–174.17) | 6; 4 (66.7%) | 1.00 (0.61–1.62) |
| IgM | 2024-25 formulation | BQ.1 | 145.73 (56.94–373.03) | 127.06 (79.25–203.70) | 6; 3 (50.0%) | 0.87 (0.49–1.55) |
| IgM | 2024-25 formulation | Original | 254.64 (179.96–360.32) | 259.52 (179.08–376.10) | 6; 3 (50.0%) | 1.02 (0.67–1.55) |
| IgM | 2024-25 formulation | XBB.1 | 148.68 (53.98–409.51) | 125.09 (81.39–192.24) | 6; 4 (66.7%) | 0.84 (0.39–1.83) |

| Antibody | Vaccine formulation | Antigen | Pre mean inhibition (95% CI) | Post mean inhibition (95% CI) | Samples (n) | Paired change |
| --- | --- | --- | --- | --- | --- | --- |
| ACE2 | Monovalent booster | Original | 97.8 | 95.3 (89.6–101.0) | Pre 1; Post 16 |  |
| ACE2 | Monovalent booster | BA.1 | 15.4 | 66.6 (37.6–95.5) | Pre 1; Post 7 |  |
| ACE2 | Monovalent booster | BA.2.75 | 51.2 | 68.0 (50.4–85.5) | Pre 1; Post 15 |  |
| ACE2 | Monovalent booster | BA.4/BA.5 | 99.0 | 78.3 (63.3–93.3) | Pre 1; Post 16 |  |
| ACE2 | Monovalent booster | BF.7 | 98.6 | 77.5 (50.5–104.5) | Pre 1; Post 7 |  |
| ACE2 | Monovalent booster | BQ.1 | 93.1 | 63.6 (32.5–94.6) | Pre 1; Post 7 |  |
| ACE2 | Monovalent booster | BN.1 | 97.5 | 78.2 (52.4–104.0) | Pre 1; Post 7 |  |
| ACE2 | Monovalent booster | XBB.1 | 85.9 | 68.2 (42.1–94.3) | Pre 1; Post 8 |  |
| ACE2 | Bivalent booster | Original | 92.6 (82.9–102.2) | 89.0 (67.8–110.2) | Pre 5; Post 8 |  |
| ACE2 | Bivalent booster | BA.4/BA.5 | 76.0 (40.4–111.5) | 78.5 (54.4–102.5) | Pre 5; Post 8 |  |
| ACE2 | Bivalent booster | XBB.1 | 66.0 (25.5–106.5) | 81.3 (58.0–104.7) | Pre 5; Post 8 |  |
| ACE2 | Bivalent booster | FL.1.5 | 31.4 (6.1–56.6) | 77.9 (55.4–100.4) | Pre 5; Post 8 |  |
| ACE2 | Bivalent booster | EG.5.1 | 53.2 (12.9–93.5) | 72.2 (44.6–99.9) | Pre 5; Post 8 |  |
| ACE2 | Bivalent booster | BA.2.86 | 57.4 (33.1–81.6) | 77.3 (55.4–99.1) | Pre 5; Post 8 |  |
| ACE2 | 2023-24 formulation | Original | 79.6 (62.8–96.4) | 95.5 (90.3–100.8) | Pre 11; Post 12 |  |
| ACE2 | 2023-24 formulation | BA.4/BA.5 | 65.8 (42.0–89.6) | 91.9 (85.6–98.2) | Pre 11; Post 12 |  |
| ACE2 | 2023-24 formulation | XBB.1 | 54.6 (31.6–77.7) | 88.4 (79.3–97.5) | Pre 11; Post 12 |  |
| ACE2 | 2023-24 formulation | FL.1.5 | 42.5 (19.5–65.6) | 86.8 (78.3–95.2) | Pre 11; Post 12 |  |
| ACE2 | 2023-24 formulation | EG.5.1 | 47.8 (23.2–72.4) | 86.7 (76.7–96.6) | Pre 11; Post 12 |  |
| ACE2 | 2023-24 formulation | JD.1.1 | 46.5 (26.9–66.1) | 79.8 (66.6–93.0) | Pre 11; Post 12 |  |
| ACE2 | 2023-24 formulation | HV.1 | 47.6 (26.5–68.6) | 81.5 (68.6–94.5) | Pre 11; Post 12 |  |
| ACE2 | 2023-24 formulation | BA.2.86 | 49.1 (25.5–72.7) | 80.9 (68.0–93.8) | Pre 11; Post 12 |  |
| ACE2 | 2023-24 formulation | JN.1 | 44.5 (24.7–64.3) | 76.8 (62.4–91.2) | Pre 11; Post 12 |  |
| ACE2 | 2024-25 formulation | Original | 95.4 (91.9–98.9) | 98.1 (96.6–99.6) | Pre 9; Post 7 |  |
| ACE2 | 2024-25 formulation | BA.4/BA.5 | 86.0 (75.9–96.1) | 95.0 (92.0–98.0) | Pre 9; Post 7 |  |
| ACE2 | 2024-25 formulation | XBB.1 | 84.6 (68.0–101.1) | 86.7 (74.7–98.7) | Pre 6; Post 5 |  |
| ACE2 | 2024-25 formulation | FL.1.5 | 83.8 (70.2–97.4) | 84.7 (68.0–101.4) | Pre 6; Post 5 |  |
| ACE2 | 2024-25 formulation | EG.5.1 | 79.7 (62.0–97.5) | 83.0 (65.7–100.4) | Pre 6; Post 5 |  |
| ACE2 | 2024-25 formulation | JD.1.1 | 73.1 (53.6–92.6) | 77.3 (55.1–99.4) | Pre 6; Post 5 |  |
| ACE2 | 2024-25 formulation | HV.1 | 73.3 (50.8–95.9) | 82.8 (65.8–99.8) | Pre 6; Post 5 |  |
| ACE2 | 2024-25 formulation | BA.2.86 | 78.2 (58.0–98.4) | 82.4 (66.6–98.2) | Pre 6; Post 5 |  |
| ACE2 | 2024-25 formulation | JN.1 | 70.0 (44.2–95.9) | 81.0 (63.0–98.9) | Pre 6; Post 5 |  |

Table S9: Power-law waning model coefficients, predicted antibody levels at 15, 180, and 300 days following vaccination, by vaccine formulation

| **Antibody** | **Vaccine formulation** | **Specimens, n** | **Participants, n** | **β (95% CI)** | **P value** | **Day 15 (95% CI)** | **Day 180 (95% CI)** | **Day 300 (95% CI)** |
| --- | --- | --- | --- | --- | --- | --- | --- | --- |
| IgG | Monovalent booster | 29 | 12 | −1.03 (−1.51 to −0.55) | <0.001 | 1,185,664 (562,300–2,500,087) | 91,191 (44,183–188,209) | 53,820 (21,170–136,825) |
|  | Bivalent booster | 34 | 16 | −1.00 (−1.71 to −0.30) | 0.007 | 414,281 (104,212–1,646,922) | 34,283 (11,456–102,597) | 20,541 (5,330–79,152) |
|  | 2023–24 formulation | 22 | 13 | 0.83 (−0.38 to 2.04) | 0.168 | 12,704 (1,421–113,569) | 99,742 (16,562–600,695) | 152,354 (15,788–1,470,245) |
|  | 2024–25 formulation | 8 | 6 | 0.02 (−0.76 to 0.79) | 0.956 | 272,393 (105,162–705,556) | 285,146 (67,589–1,202,971) | 287,840 (47,676–1,737,831) |
| ACE2 | Monovalent booster | 29 | 12 | −9.40 (−15.55 to −3.26) | 0.004 | 99.8 (86.0–113.6) | 66.1 (52.7–79.5) | 59.2 (41.9–76.5) |
|  | Bivalent booster | 34 | 16 | −5.57 (−11.82 to 0.69) | 0.079 | 91.1 (73.5–108.8) | 71.2 (57.2–85.2) | 67.1 (49.8–84.3) |
|  | 2023–24 formulation | 19 | 12 | 0.58 (−12.87 to 14.03) | 0.929 | 72.3 (42.0–102.5) | 74.3 (43.5–105.1) | 74.8 (35.6–113.9) |
| IgM | Monovalent booster | 29 | 12 | −0.60 (−0.97 to −0.23) | 0.002 | 773 (436–1,372) | 174 (100–304) | 128 (63–263) |
|  | Bivalent booster | 34 | 16 | −0.16 (−0.37 to 0.05) | 0.128 | 295 (195–445) | 197 (142–273) | 181 (121–271) |
|  | 2023–24 formulation | 22 | 13 | −0.28 (−0.73 to 0.17) | 0.212 | 231 (102–521) | 116 (59–225) | 100 (43–233) |
|  | 2024–25 formulation | 8 | 6 | −0.86 (−1.84 to 0.12) | 0.076 | 316 (95–1,050) | 37 (6–230) | 24 (2–233) |

Table S10: Peak post-vaccination and late follow-up antibody responses across SARS-CoV-2 variants and vaccine formulations

| **Antibody** | **Vaccine formulation** | **Antigen** | **Peak (95% CI)** | **Peak n; Late n** | **Late (95% CI)** | **Paired fold change / mean change (95% CI)** |
| --- | --- | --- | --- | --- | --- | --- |
| IgG | Monovalent booster | BA.1 | 89,788 (45,666–176,539) | 17; 16 | 79,423 (40,976–153,945) | 0.64 (0.30–1.37) |
| IgG | Monovalent booster | BA.2.75 | 74,921 (38,593–145,446) | 17; 16 | 65,943 (33,835–128,522) | 0.65 (0.29–1.46) |
| IgG | Monovalent booster | BA.4.6 | 82,027 (43,649–154,150) | 17; 16 | 70,476 (35,777–138,829) | 0.64 (0.30–1.35) |
| IgG | Monovalent booster | BA.4/BA.5 | 102,028 (54,502–190,996) | 17; 16 | 89,103 (45,120–175,961) | 0.64 (0.30–1.35) |
| IgG | Monovalent booster | BF.7 | 76,925 (40,966–144,447) | 17; 16 | 65,292 (33,191–128,437) | 0.63 (0.30–1.33) |
| IgG | Monovalent booster | BN.1 | 123,988 (28,540–538,653) | 5; 5 | 35,010 (17,809–68,825) | 0.38 (0.10–1.47) |
| IgG | Monovalent booster | BQ.1 | 72,035 (39,397–131,714) | 17; 16 | 60,487 (30,599–119,570) | 0.63 (0.30–1.33) |
| IgG | Monovalent booster | Original | 411,250 (221,144–764,781) | 17; 16 | 382,741 (210,227–696,822) | 0.73 (0.35–1.49) |
| IgG | Monovalent booster | XBB.1 | 70,151 (38,040–129,368) | 17; 16 | 58,224 (29,576–114,624) | 0.64 (0.30–1.38) |
| IgG | Bivalent booster | BA.1 | 216,995 (99,368–473,864) | 15; 15 | 9,934 (1,509–65,424) | 0.04 (0.01–0.26) |
| IgG | Bivalent booster | BA.2.75 | 180,735 (81,273–401,919) | 15; 15 | 8,934 (1,288–61,973) | 0.05 (0.01–0.30) |
| IgG | Bivalent booster | BA.4.6 | 222,179 (98,604–500,627) | 15; 15 | 11,754 (1,763–78,350) | 0.05 (0.01–0.31) |
| IgG | Bivalent booster | BA.4/BA.5 | 287,779 (126,961–652,302) | 15; 15 | 15,513 (2,412–99,766) | 0.05 (0.01–0.30) |
| IgG | Bivalent booster | BF.7 | 202,624 (90,853–451,903) | 15; 15 | 9,855 (1,421–68,338) | 0.05 (0.01–0.29) |
| IgG | Bivalent booster | BQ.1 | 184,902 (83,887–407,558) | 15; 15 | 9,665 (1,485–62,879) | 0.05 (0.01–0.30) |
| IgG | Bivalent booster | Original | 842,477 (388,609–1,826,432) | 15; 15 | 33,167 (4,627–237,754) | 0.04 (0.01–0.28) |
| IgG | Bivalent booster | XBB.1 | 159,735 (74,099–344,340) | 15; 15 | 8,103 (1,240–52,950) | 0.05 (0.01–0.30) |
| IgG | 2023-24 formulation | BA.1 | 30,141 (3,908–232,486) | 12; 11 | 104,649 (53,948–202,998) | 5.02 (0.43–58.03) |
| IgG | 2023-24 formulation | BA.2.75 | 28,956 (3,391–247,228) | 12; 11 | 114,390 (59,726–219,086) | 5.81 (0.40–84.74) |
| IgG | 2023-24 formulation | BA.4.6 | 150 (104–217) | 3; 9 | 104,673 (51,774–211,619) | 494.08 (5.01–48,688.59) |
| IgG | 2023-24 formulation | BA.4/BA.5 | 46,016 (5,519–383,697) | 12; 11 | 143,861 (80,759–256,268) | 4.52 (0.37–54.58) |
| IgG | 2023-24 formulation | BF.7 | 32,086 (3,723–276,496) | 12; 11 | 113,293 (63,022–203,665) | 5.15 (0.44–60.70) |
| IgG | 2023-24 formulation | BN.1 | 142,731 (89,582–227,414) | 10; 10 | 70,619 (37,067–134,540) | 0.57 (0.27–1.20) |
| IgG | 2023-24 formulation | BQ.1 | 27,164 (3,299–223,638) | 12; 11 | 90,089 (51,953–156,218) | 4.81 (0.45–51.64) |
| IgG | 2023-24 formulation | Original | 82,710 (8,816–775,975) | 12; 11 | 323,308 (169,092–618,173) | 5.60 (0.38–83.20) |
| IgG | 2023-24 formulation | XBB.1 | 33,816 (3,732–306,394) | 12; 11 | 115,526 (60,180–221,774) | 5.29 (0.40–70.85) |
| IgG | 2024-25 formulation | BA.1 | 273,616 (161,326–464,064) | 7; 3 | 239,174 (16,365–3,495,509) | 0.69 (0.24–1.98) |
| IgG | 2024-25 formulation | BA.2.75 | 270,645 (161,862–452,539) | 7; 3 | 225,565 (12,129–4,195,016) | 0.70 (0.19–2.59) |
| IgG | 2024-25 formulation | BA.4.6 | 276,737 (208,702–366,952) | 7; 3 | 215,627 (28,678–1,621,279) | 0.73 (0.21–2.53) |
| IgG | 2024-25 formulation | BA.4/BA.5 | 361,997 (269,711–485,861) | 7; 3 | 268,249 (33,074–2,175,672) | 0.70 (0.18–2.71) |
| IgG | 2024-25 formulation | BF.7 | 259,550 (196,597–342,660) | 7; 3 | 193,424 (26,282–1,423,516) | 0.69 (0.19–2.45) |
| IgG | 2024-25 formulation | BQ.1 | 233,092 (193,657–280,556) | 7; 3 | 158,633 (33,859–743,218) | 0.66 (0.21–2.10) |
| IgG | 2024-25 formulation | Original | 694,438 (399,645–1,206,682) | 7; 3 | 712,767 (52,746–9,631,786) | 0.88 (0.25–3.18) |
| IgG | 2024-25 formulation | XBB.1 | 279,717 (177,400–441,046) | 7; 3 | 218,176 (14,867–3,201,827) | 0.67 (0.21–2.13) |
| ACE2 | Monovalent booster | BA.1 | 81.8 (39.1–124.6) | 5; 4 | 38.9 (-22.0–99.7) | -47.6 (-608.8–513.6) |
| ACE2 | Monovalent booster | BA.2.75 | 70.2 (50.1–90.4) | 14; 4 | 68.5 (21.9–115.0) | 11.4 (-123.6–146.4) |
| ACE2 | Monovalent booster | BA.4/BA.5 | 75.3 (57.0–93.5) | 14; 7 | 81.0 (49.3–112.7) | 5.1 (-32.4–42.6) |
| ACE2 | Monovalent booster | BF.7 | 81.5 (34.3–128.6) | 5; 4 | 80.0 (33.5–126.5) | -5.0 (-33.2–23.1) |
| ACE2 | Monovalent booster | BN.1 | 82.8 (38.8–126.8) | 5; 4 | 60.8 (-6.1–127.6) | -43.8 (-569.6–481.9) |
| ACE2 | Monovalent booster | BQ.1 | 65.2 (8.8–121.5) | 5; 4 | 55.6 (-10.4–121.6) | -12.4 (-928.8–904.1) |
| ACE2 | Monovalent booster | Original | 94.8 (87.6–102.0) | 14; 7 | 92.1 (82.9–101.3) | 0.7 (-14.9–16.3) |
| ACE2 | Monovalent booster | XBB.1 | 63.9 (7.1–120.8) | 5; 7 | 48.1 (7.2–89.0) | -39.6 (-93.6–14.4) |
| ACE2 | Bivalent booster | BA.2.86 | 77.3 (50.9–103.7) | 8; 12 | 58.4 (32.5–84.3) | -6.6 (-64.4–51.1) |
| ACE2 | Bivalent booster | BA.4/BA.5 | 78.5 (49.4–107.5) | 8; 12 | 72.8 (47.5–98.0) | 11.9 (-27.5–51.3) |
| ACE2 | Bivalent booster | EG.5.1 | 72.2 (38.8–105.6) | 8; 12 | 56.9 (30.7–83.0) | 1.9 (-50.1–53.9) |
| ACE2 | Bivalent booster | FL.1.5 | 77.9 (50.7–105.0) | 8; 12 | 52.2 (26.5–77.8) | -9.4 (-72.0–53.3) |
| ACE2 | Bivalent booster | Original | 89.0 (63.4–114.6) | 8; 12 | 82.2 (64.3–100.0) | 1.7 (-50.2–53.5) |
| ACE2 | Bivalent booster | XBB.1 | 81.3 (53.1–109.5) | 8; 12 | 63.6 (38.5–88.7) | 0.8 (-50.3–51.9) |
| ACE2 | 2023-24 formulation | BA.2.86 | 80.9 (66.4–95.4) | 12; 8 | 62.4 (32.9–91.9) | -11.4 (-37.8–14.9) |
| ACE2 | 2023-24 formulation | BA.4/BA.5 | 91.9 (84.9–99.0) | 12; 11 | 64.9 (41.8–88.0) | -27.2 (-54.1–-0.3) |
| ACE2 | 2023-24 formulation | EG.5.1 | 86.7 (75.5–97.9) | 12; 8 | 64.7 (32.6–96.8) | -16.8 (-53.2–19.6) |
| ACE2 | 2023-24 formulation | FL.1.5 | 86.8 (77.3–96.3) | 12; 8 | 68.8 (37.9–99.7) | -15.1 (-49.4–19.1) |
| ACE2 | 2023-24 formulation | HV.1 | 81.5 (67.0–96.1) | 12; 8 | 60.1 (27.3–93.0) | -13.9 (-44.1–16.3) |
| ACE2 | 2023-24 formulation | JD.1.1 | 79.8 (65.0–94.7) | 12; 8 | 58.6 (27.4–89.8) | -13.8 (-40.2–12.7) |
| ACE2 | 2023-24 formulation | JN.1 | 76.8 (60.6–92.9) | 12; 8 | 55.1 (21.3–88.8) | -14.6 (-48.8–19.7) |
| ACE2 | 2023-24 formulation | Original | 95.5 (89.6–101.5) | 12; 11 | 80.9 (60.3–101.6) | -15.9 (-33.1–1.4) |
| ACE2 | 2023-24 formulation | XBB.1 | 88.4 (78.2–98.6) | 12; 8 | 66.1 (31.7–100.5) | -17.6 (-55.0–19.8) |
| IgM | Monovalent booster | BA.1 | 158 (105–238) | 17; 16 | 150 (104–215) | 0.82 (0.62–1.09) |
| IgM | Monovalent booster | BA.2.75 | 133 (82–215) | 17; 16 | 130 (84–201) | 0.86 (0.67–1.10) |
| IgM | Monovalent booster | BA.4.6 | 157 (111–222) | 17; 16 | 128 (94–175) | 0.82 (0.59–1.15) |
| IgM | Monovalent booster | BA.4/BA.5 | 216 (151–309) | 17; 16 | 179 (129–248) | 0.85 (0.60–1.21) |
| IgM | Monovalent booster | BF.7 | 134 (93–192) | 17; 16 | 108 (78–151) | 0.80 (0.58–1.10) |
| IgM | Monovalent booster | BQ.1 | 144 (102–204) | 17; 16 | 121 (87–169) | 0.88 (0.62–1.24) |
| IgM | Monovalent booster | Original | 507 (329–780) | 17; 16 | 396 (252–622) | 0.83 (0.57–1.21) |
| IgM | Monovalent booster | XBB.1 | 118 (76–182) | 17; 16 | 115 (79–168) | 0.85 (0.62–1.17) |
| IgM | Bivalent booster | BA.1 | 260 (192–352) | 15; 15 | 163 (114–232) | 0.61 (0.35–1.09) |
| IgM | Bivalent booster | BA.2.75 | 219 (152–316) | 15; 15 | 169 (107–267) | 0.77 (0.35–1.67) |
| IgM | Bivalent booster | BA.4.6 | 240 (165–349) | 15; 15 | 203 (126–328) | 0.82 (0.43–1.57) |
| IgM | Bivalent booster | BA.4/BA.5 | 353 (237–527) | 15; 15 | 300 (183–492) | 0.85 (0.43–1.67) |
| IgM | Bivalent booster | BF.7 | 195 (136–280) | 15; 15 | 161 (103–252) | 0.80 (0.42–1.50) |
| IgM | Bivalent booster | BN.1 | — | 0; 2 | 89 (1–6,362) | — |
| IgM | Bivalent booster | BQ.1 | 229 (155–337) | 15; 15 | 189 (111–321) | 0.81 (0.38–1.72) |
| IgM | Bivalent booster | Original | 475 (334–676) | 15; 15 | 314 (230–429) | 0.69 (0.44–1.08) |
| IgM | Bivalent booster | XBB.1 | 203 (137–301) | 15; 15 | 159 (89–286) | 0.83 (0.35–1.95) |
| IgM | 2023-24 formulation | BA.1 | 238 (122–463) | 12; 11 | 121 (86–169) | 0.70 (0.51–0.95) |
| IgM | 2023-24 formulation | BA.2.75 | 195 (92–413) | 12; 11 | 101 (71–144) | 0.75 (0.53–1.04) |
| IgM | 2023-24 formulation | BA.4.6 | 150 (104–217) | 3; 9 | 92 (56–151) | 0.57 (0.25–1.30) |
| IgM | 2023-24 formulation | BA.4/BA.5 | 271 (127–580) | 12; 11 | 127 (87–184) | 0.67 (0.51–0.87) |
| IgM | 2023-24 formulation | BF.7 | 180 (83–389) | 12; 11 | 84 (58–122) | 0.68 (0.51–0.90) |
| IgM | 2023-24 formulation | BN.1 | 270 (93–787) | 10; 10 | 103 (69–153) | 0.63 (0.40–1.00) |
| IgM | 2023-24 formulation | BQ.1 | 196 (91–423) | 12; 11 | 92 (63–136) | 0.69 (0.51–0.92) |
| IgM | 2023-24 formulation | Original | 290 (213–396) | 12; 11 | 221 (154–315) | 0.77 (0.60–0.99) |
| IgM | 2023-24 formulation | XBB.1 | 231 (102–521) | 12; 11 | 99 (70–139) | 0.66 (0.50–0.89) |
| IgM | 2024-25 formulation | BA.1 | 199 (79–498) | 7; 3 | 197 (5–8,193) | 0.44 (0.10–1.92) |
| IgM | 2024-25 formulation | BA.2.75 | 160 (64–400) | 7; 3 | 129 (16–1,048) | 0.38 (0.07–2.06) |
| IgM | 2024-25 formulation | BA.4.6 | 187 (75–468) | 7; 3 | 152 (126–182) | 0.42 (0.02–7.29) |
| IgM | 2024-25 formulation | BA.4/BA.5 | 270 (110–664) | 7; 3 | 223 (199–248) | 0.41 (0.04–4.62) |
| IgM | 2024-25 formulation | BF.7 | 153 (72–325) | 7; 3 | 130 (113–149) | 0.51 (0.05–5.11) |
| IgM | 2024-25 formulation | BQ.1 | 181 (67–492) | 7; 3 | 148 (116–189) | 0.39 (0.02–7.02) |
| IgM | 2024-25 formulation | Original | 287 (181–458) | 7; 3 | 290 (118–710) | 0.71 (0.19–2.75) |
| IgM | 2024-25 formulation | XBB.1 | 188 (63–561) | 7; 3 | 100 (12–808) | 0.23 (0.01–4.00) |
